# Cell-type specific cis-eQTLs in eight brain cell-types identifies novel risk genes for human brain disorders

**DOI:** 10.1101/2021.10.09.21264604

**Authors:** Julien Bryois, Daniela Calini, Will Macnair, Lynette Foo, Eduard Urich, Ward Ortmann, Victor Alejandro Iglesias, Suresh Selvaraj, Erik Nutma, Manuel Marzin, Sandra Amor, Anna Williams, Gonçalo Castelo-Branco, Vilas Menon, Philip De Jager, Dheeraj Malhotra

## Abstract

Most expression quantitative trait loci (eQTL) studies to date have been performed in heterogeneous brain tissues as opposed to specific cell types. To investigate the genetics of gene expression in adult human cell types from the central nervous system (CNS), we performed an eQTL analysis using single nuclei RNA-seq from 196 individuals in eight CNS cell types. We identified 6108 eGenes, a substantial fraction (43%, 2620 out of 6108) of which show cell-type specific effects, with strongest effects in microglia. Integration of CNS cell-type eQTLs with GWAS revealed novel relationships between expression and disease risk for neuropsychiatric and neurodegenerative diseases. For most GWAS loci, a single gene colocalized in a single cell type providing new clues into disease etiology. Our findings demonstrate substantial contrast in genetic regulation of gene expression among CNS cell types and reveal genetic mechanisms by which disease risk genes influence neurological disorders.

## Introduction

Most genetic associations from genome-wide association studies (GWAS) of brain disorders lie within non-coding regions of the genome, challenging the identification of risk genes ^1^, and CNS cell types in which these risk variants regulate gene expression. Expression quantitative trait loci (eQTLs) (i.e. genomic regions that explain variation in gene expression levels) have become a powerful tool to uncover the molecular underpinnings of variants associated with complex traits and diseases in non-coding regions of the genome ^2–4^.

Importantly, eQTL relationships are highly dependent upon cell type ^5^, cell states and developmental stage of the human brain ^6–8^, consistent with recent transcriptomic studies that show prominent temporal and cell type specific changes in expression ^9,10^. However, most prior eQTL studies were done using bulk human brain tissues and have been partially successful in prioritizing disease risk genes by integrating GWAS results with tissue-level eQTLs. A few recent studies have investigated eQTLs in specific CNS cell types, for example, cis-eQTLs in dopaminergic neurons derived from induced pluripotent stem cells ^11^, and in sorted primary human microglia ^12,13^. To dissect the functional genetic variation of late-onset neuropsychiatric and neurological diseases, we leveraged single-nucleus gene expression analysis from 196 adult human brain tissues (both cortical grey matter and deep white matter) to perform a systematic eQTL analysis in all major adult human CNS cell types.

Here, we present the first single-cell based map of eQTLs in eight human CNS cell-types. We show substantial cell-type specific effects in the genetic control of gene expression. Integration of cell-type eQTLs with GWAS shows that, at most GWAS loci, a single gene colocalizes in a single CNS cell-type thereby not only providing insights into disease-relevant genes but also identifying new putative mechanisms of risk genes that have been missed by bulk tissue-level analyses.

## Results

### Robust identification, fine mapping and functional characterization of brain cell type cis-eQTLs

Our goal was to identify genetic variants regulating gene expression in CNS cell types. Therefore, we performed single nuclei RNA-seq and genotyping in 246 human brain samples from 123 independent individuals (**Table S1**). We integrated our dataset with 127 human brain samples from published single-cell transcriptomic studies that were previously genotyped ^14–16^, resulting in a total of 373 brain samples from 215 individuals. After quality control, normalization, sample integration and genotype imputation (**Online Methods**), we obtained gene expression data for 7208-10846 genes (protein coding and non-coding) and genotypes for 5.3 million SNPs in 196 individuals for 8 major CNS cell types (**Figure 1**) expressing clear canonical markers of cell type identity (**Figure S1**).

**Figure 1:**
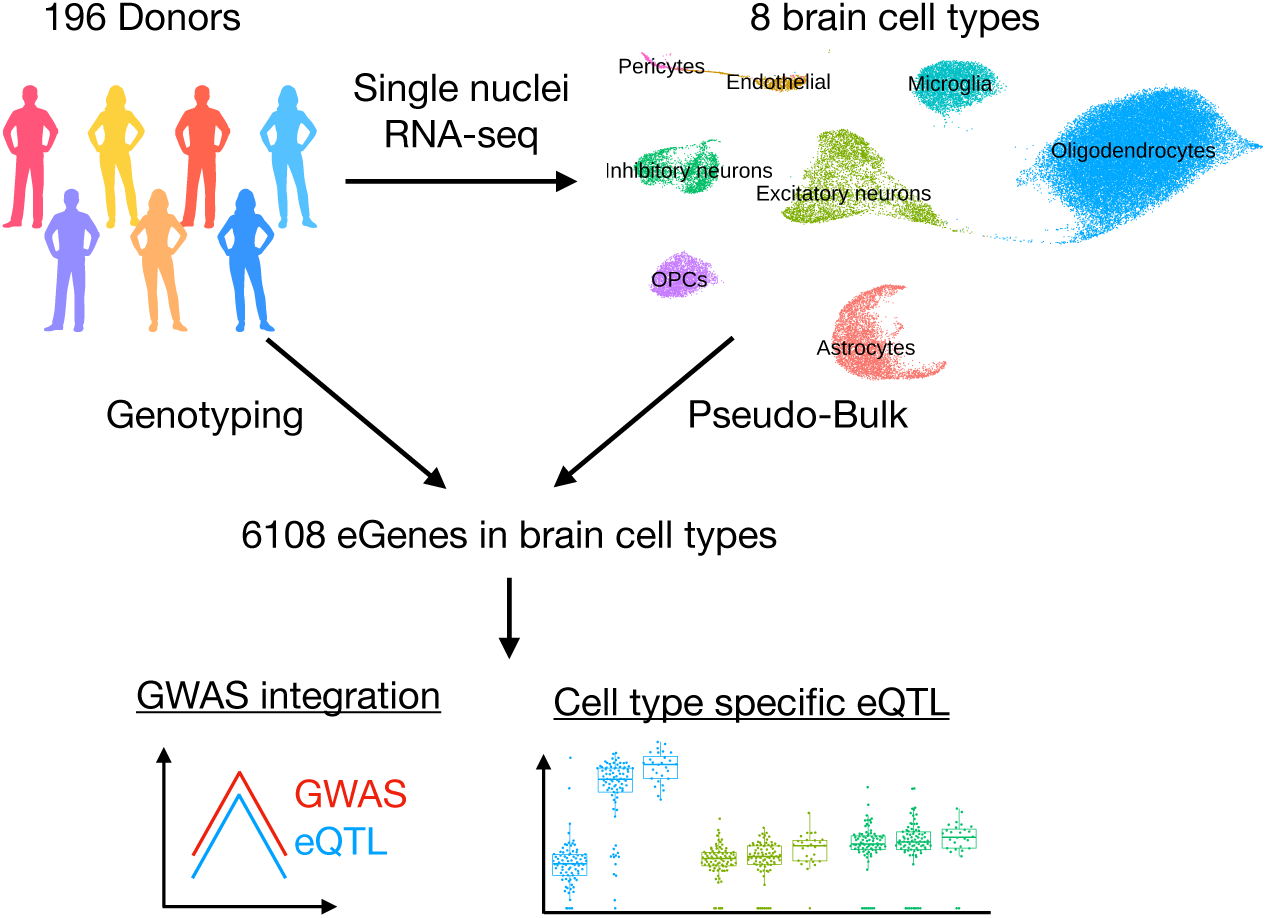
Study summary We performed single nuclei RNA-seq on brain samples from 196 genotyped donors. We mapped cis-eQTLs for 8 major brain cell types and identified a total of 6108 cis-eQTL genes. We identified cell type specific genetic effects and leveraged our results to identify risk genes for brain disorders.

We identified cis-eQTLs by testing all SNPs within a 1 megabase (MB) window surrounding the transcription start site (TSS) of each expressed gene while adjusting for known covariates (study, disease status) and inferred covariates (genotype first principal components (PCs), expression first PCs) (**Online Methods**). We discovered 6108 genes with a cis-eQTL at a 5% false discovery rate (FDR) across the 8 different CNS cell types (**Figure 2A, Table S2**). This number represents only a small fraction of the potential cis-eQTL discoveries as we estimate that at least 10-50% of the tested genes have an eQTL across the different cell types (**Figure S2, Online Methods**). Most cis-eQTLs replicated in a large tissue-level cortical eQTL study (Metabrain ^17^) with 72.1-82.3% of the SNP-gene pairs having a pvalue <0.05 in this larger study (**Figure S3A**). Cis-eQTLs that did not replicate (p>0.05) affected more constrained genes (**Figure S3B**) and were located further from the TSS than replicating cis-eQTLs (**Figure S3C**). Neuron cis-eQTLs replicated at a higher rate (80.1%, pi1=86%) than glia cis-eQTL (75%, pi1=77%) (**Figure 2C**), possibly because some glial cell types are less prevalent than neurons in the cortex. The number of detected eQTLs varied significantly between cell types (**Figure 2A**) (e.g. 2114 eQTL in excitatory neurons but only 23 in pericytes) and showed high correlation with the total number of nuclei belonging to the cell type (**Figure 2B**). This suggests that more nuclei allow for a better quantification of gene expression and, ultimately, the discovery of more eQTLs.

**Figure 2:**
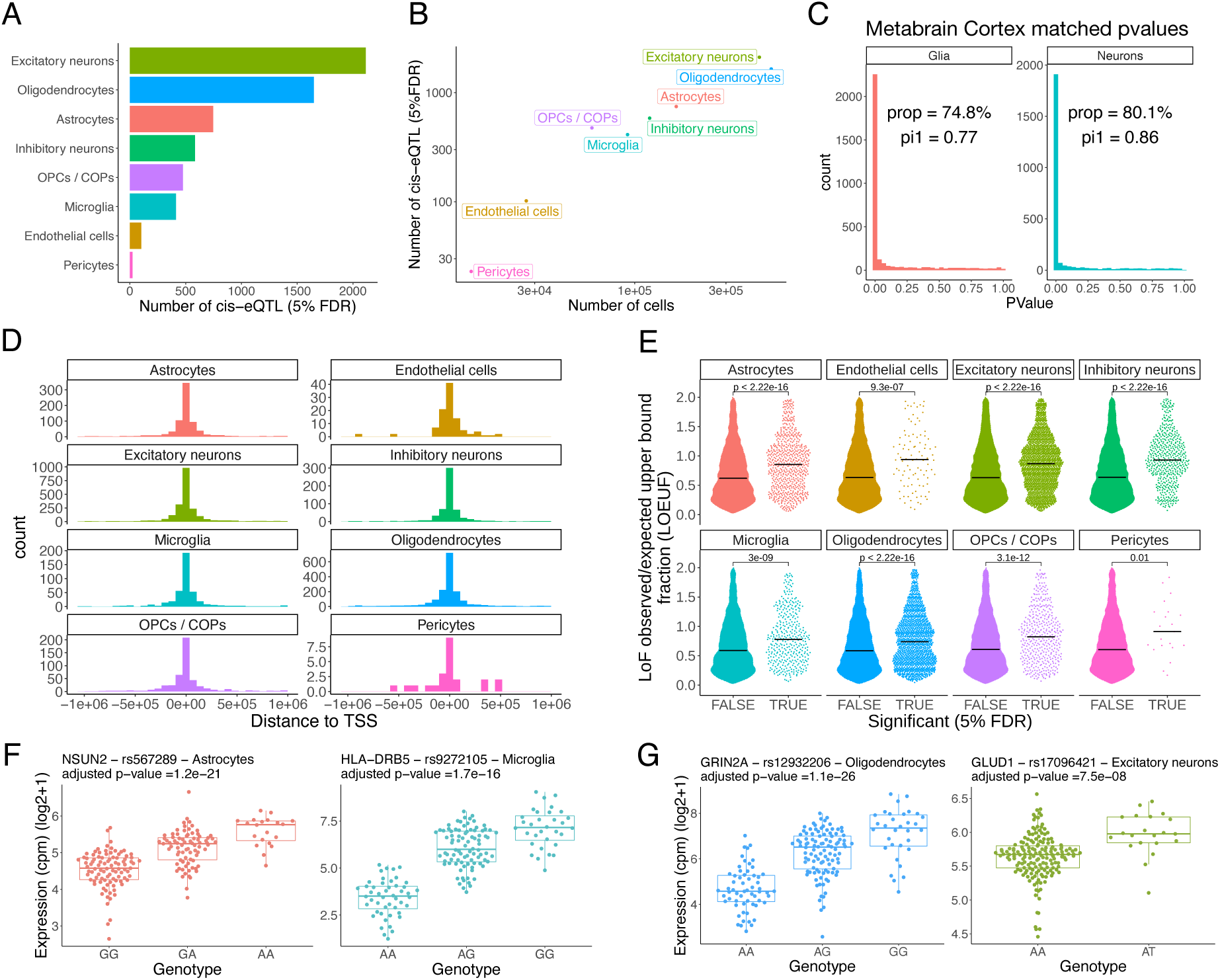
Cis-eQTL discoveries **A**) Number of cis-eQTLs per cell types (5% FDR). **B**) Number of cis-eQTLs (5% FDR) versus number of single nuclei belonging to the cell types. **C**) Replication pvalues for each SNP-gene pairs of our cis-eQTL discoveries (aggregated for glia and neurons) in a large cortical eQTL study ^17^. **D**) Enrichment of the discovered cis-eQTLs around the TSS. **E**) LOEUF scores ^18^ for genes with a cis-eQTL (5% FDR) and genes without a cis-eQTL (constrained genes have low LOEUF score, not constrained genes have high LOEUF score). The black horizontal bars indicate the medians. **F)** Examples of cis-eQTLs in astrocytes and microglia. **G)** Examples of cis-eQTLs with a fine-mapped SNPs (causal probability of 0.94 for GRIN2A and 0.87 for GLUD1).

As expected, cis-eQTLs were enriched around the TSS (**Figure 2D**) and more frequently found upstream of the gene or within the gene body than downstream of the gene (**Figure S4**) (2679 in gene body, 2156 upstream, 1273 downstream). We found that 48-59% of the top cis-eQTL SNP affected the closest gene (depending on the cell type) (**Figure S5, Online Methods**). Genes with a cis-eQTL were found to be less constrained than genes without a cis-eQTL (**Figure 2E**), suggesting that less constrained genes are also more tolerant to variability in gene expression levels. Genes with an eQTL in glial classes had on average higher expression levels than genes without an eQTL (Wilcoxon pvalue=5*10^−9^), while the opposite was true for neurons (Wilcoxon pvalue=1*10^−5^) (**Figure S6**). Altogether, these results suggest that cell type level cis-eQTLs have similar properties as tissue level cis-eQTLs ^2^.

We next investigated whether any of the genes with a cis-eQTL had additional independent cis-eQTLs. We found that 126 genes had a secondary cis-eQTL, with the majority being discovered in excitatory neurons and oligodendrocytes (112 genes) (**Figure S7A**,**B**). Independent cis-eQTLs were enriched around the TSS of the target gene but located at a larger distance from the TSS than the main cis-eQTLs (mean distance = 243kb vs 113kb, Wilcoxon p=2*10-9) (**Figure S7C**,**D**). Larger sample sizes will be necessary to identify more independent cis-eQTLs for cell types from the CNS.

We also performed fine-mapping to identify putative causal SNPs for our cis-eQTLs (**Table S3, Online Methods**). As expected, fine-mapped SNPs with higher probability of being causal were more likely to overlap epigenomic marks ^19,20^ (**Figure S8A**). Overall, we fine-mapped 413 cis-eQTLs (causal probability>50%) (**Figure S8B**). Examples of fine-mapped SNPs with high causal probabilities include rs12932206 as the likely causal SNPs for the oligodendrocyte GRIN2A eQTL (probability=0.94) (**Figure 2G**), a gene associated with schizophrenia ^21^, and rs17096421 as the likely causal SNP for the GLUD1 excitatory neuron eQTL (probability=0.87) (**Figure 2G**), a gene with an important role in inhibitory synapse formation^22^.

We reasoned that CNS cell-type eQTLs could overlap with CNS cell-type specific regulatory regions defined by snATAC-seq ^20^ and sought to test this as an external validation (**Figure 3A, Online Methods**). We found that ATAC-seq peaks specific to glial cell types (astrocytes, microglia, oligodendrocytes and OPCs / COPs) were enriched around the cis-eQTLs discovered in the same cell types but not in the other cell types, suggesting that the discovered glial eQTLs fall in regions of the genome functionally relevant to cell-type specific gene regulation. Surprisingly, we did not observe enrichment of neuronal cis-eQTLs in neuron-specific ATAC-seq peaks, neuron-specific CHIP-seq marks ^19^ (H3K4me3 and H3K27ac) (**Figure S9**) and sorted nuclei bulk ATAC-seq ^23^ (**Figure S10**). The lack of enrichment of neuronal specific regulatory regions around neuronal cis-eQTLs was robust to multiple attempts at falsification (**Online Methods**), suggesting that the discovered neuronal cis-eQTLs might be less cell-type specific than glial cis-eQTLs.

**Figure 3:**
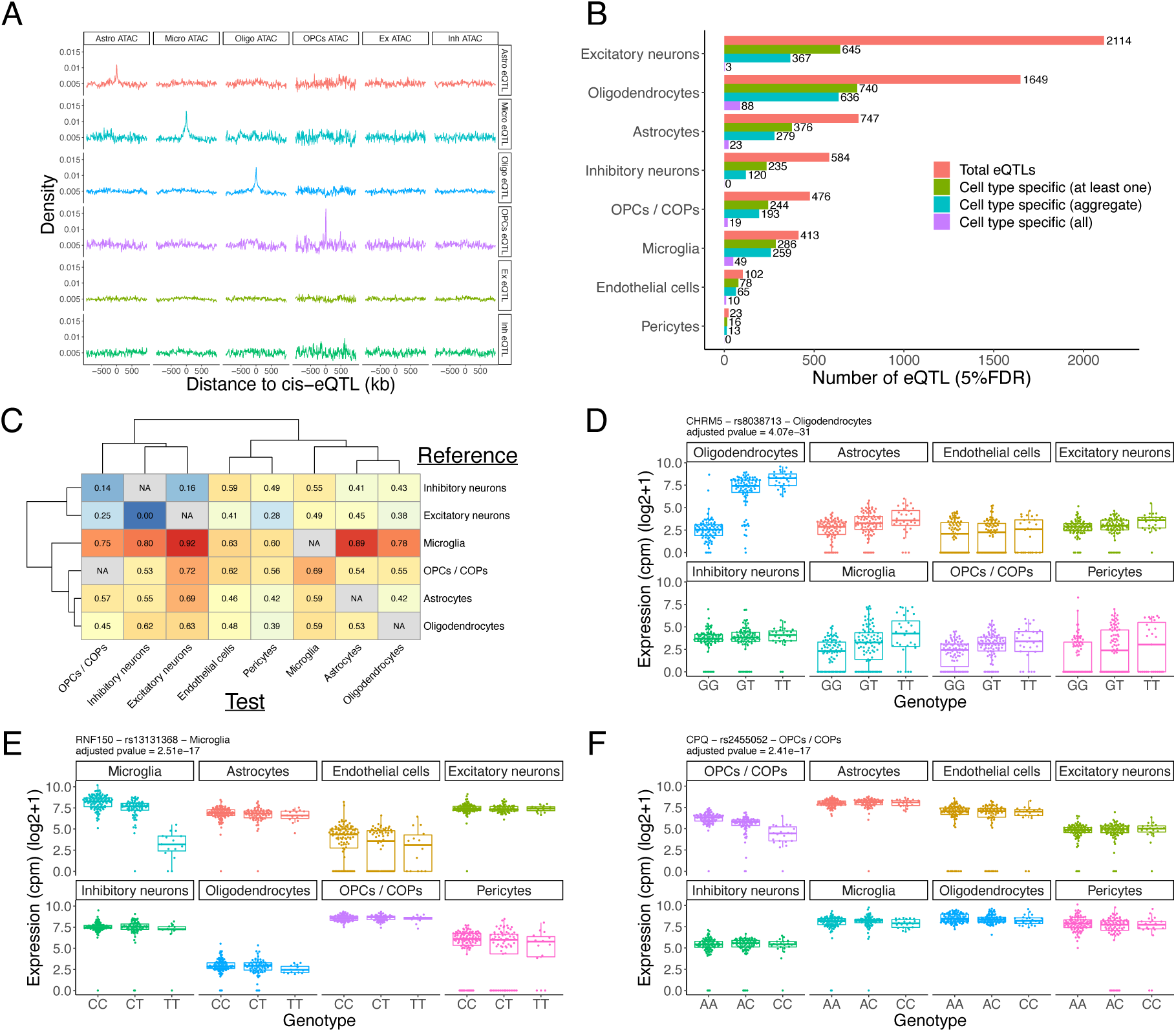
Cell type specific genetic effects on gene expression **A**) Enrichment of cell type specific ATAC-seq peaks (derived from snATAC-seq ^20^) around our cis-eQTLs. **B**) Number of significant cell type specific genetic effects (5% FDR). “Cell type specific (at least one)” shows the number of cis-eQTLs that have a different genetic effect for the same SNP-gene pair in at least one cell type. “Cell type specific (all)” shows the number of cis-eQTLs that have a different genetic for the same SNP-gene pair in all cell types, while “Cell type specific (aggregate)” shows the number of genes that have a different genetic effect in the discovered cell types than the aggregate genetic effect of all other cell types. **C**) Estimates of the proportions of cis-eQTLs that have a different genetic effect in another cell type. Estimates were computed on the interaction pvalue distributions using the pi1 statistic^25^. **D**) Example of a cell type specific cis-eQTL for CHRM5 in oligodendrocytes, **E)** RNF150 in microglia, **F)** CPQ in OPCs / COPs. Each dot represents an individual. The displayed pvalues are the adjusted interaction pvalues testing whether the genetic effect in the discovered cell type (top left cell type) is different from the genetic effects in all other cell types.

### Cell-type specific eQTL effects

We used a negative binomial mixed model to investigate how many of the 6108 cis-eQTLs had a significantly different effect size in the test cell type compared to a) the average effect size across the 7 other cell types, b) at least one of the 7 cell types, and c) all other 7 cell types (**Online Methods**). We found 1932, 2620 and 192 genes (5% FDR) respectively for the above-mentioned comparisons (**Figure 3B, Table S4)**. eGenes from a) and b) were positionally enriched around TSS compared to shared eQTLs (**Figure S11A, B**). Among all cell-type specific eGenes, excitatory neuron specific eGenes were significantly less constrained compared to the shared neuronal eQTL genes (**Figure S12A, B**). Interestingly, the 192 cell-type specific eGenes whose effect size is different than all other cell types are more constrained (**Figure S12C**), with the strongest evidence for microglia specific cis-eQTLs genes (which were also enriched in Alzheimer’s genetic associations (**Figure S13**)). Examples of these genes include CHRM5, GRIN2A and MSRA in oligodendrocytes (**Figure 3D, Figure S14**), or RNF150 in microglia and CPQ in OPCs / COPs (**Figure 3E**,**F**). GRIN2A is associated with schizophrenia ^21^, and MSRA was shown to protect dopaminergic neurons from cell death ^24^.

We used the pi1 statistic ^25^ to estimate the proportion of eQTLs with cell type specific effects for all pairwise comparisons across cell types. We found that cell-type specific genetic effects ranged from 0-92% depending on the cell types compared. For example, all eQTLs detected in excitatory neurons have similar effect sizes in inhibitory neurons (pi1=0), while 92% of the microglia eQTLs have a different effect size in excitatory neurons (pi1=0.92) (**Figure 3C** and **Figure S15**). Oligodendrocytes, OPCs / COPs, and astrocytes clustered together based on the estimated proportion of genetic effects that are cell type specific (**Figure 3C**). Similarly, neurons clustered together with an estimate that only 0-16% of the eQTLs have a different genetic effect in the other neuronal type. Microglia showed strongest evidence for cell-type specific genetic effects with an estimate that 60-92% of the discovered cis-eQTLs have a different genetic effect in the other cell types, reflecting its unique developmental origin. Altogether, these results suggest that there are substantial differences in the genetic regulation of gene expression across brain cell types, which might be relevant for brain disorders.

### CNS cell type eQTLs mediate neurological disease association

We used Coloc ^3^ to investigate whether genetic variants associated with risk of Alzheimer’s disease ^4^, Parkinson’s disease ^26,27^, schizophrenia ^28^ and multiple sclerosis ^29^ potentially act through CNS cell-type cis-eQTLs (**Table S5**). Colocalization analysis identified risk genes in CNS cell types at 13-38% of the GWAS loci (at a posterior probability (PP) > 0.7) across the four different disorders (**Figure S16**). Notably, we found that 75-93% of colocalized loci contained a single colocalized gene (**Figure 4A**) that usually occurred in a single cell type (**Figure 4A**,**B**). This suggests that disease risk at a given GWAS locus is usually mediated by a single gene acting in a specific cell type.

**Figure 4:**
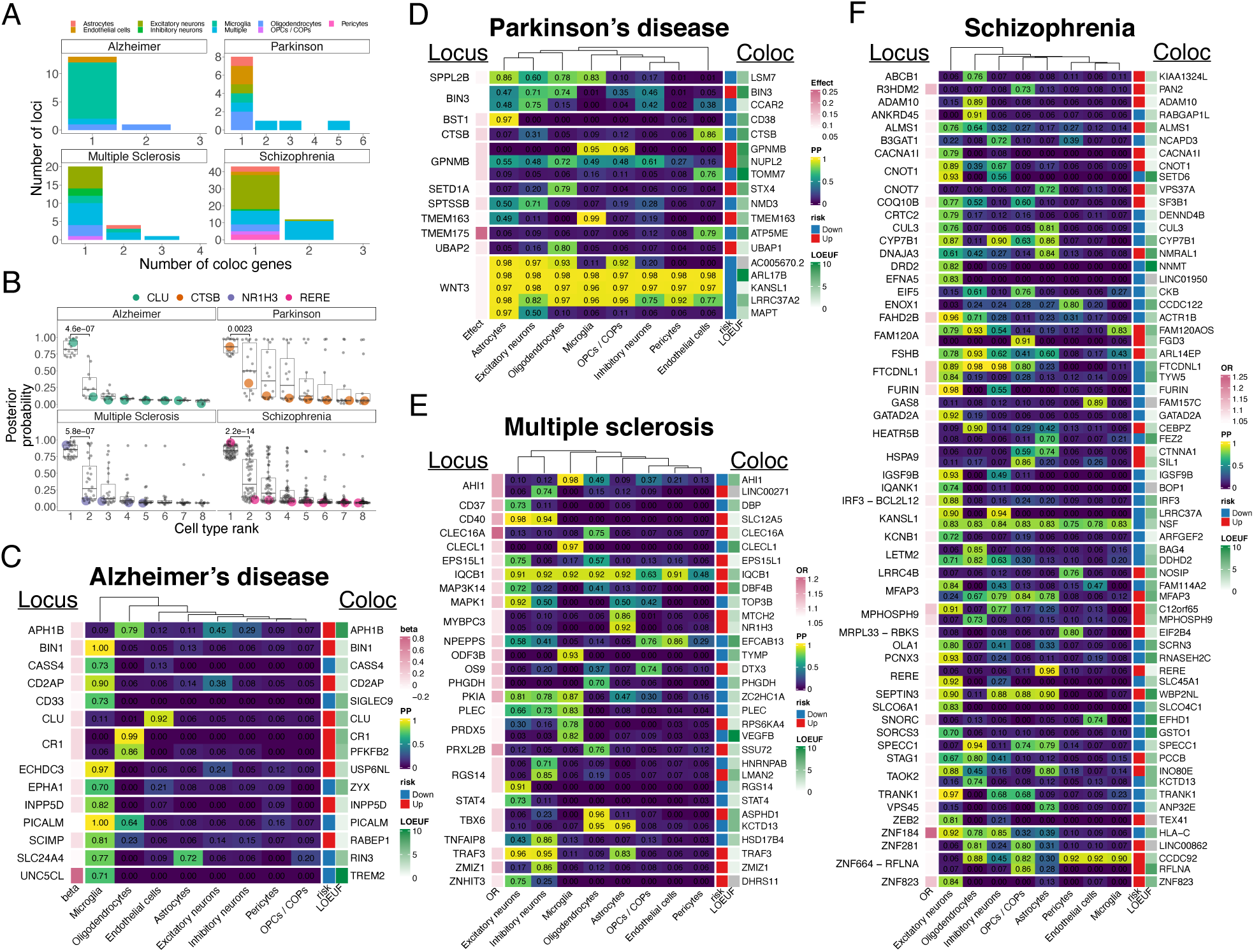
Colocalization results **A)** Number of colocalized genes per loci. At most loci a single gene colocalizes in a single cell type. **B)** Posterior probability (PP) of colocalization for colocalized genes (PP>0.7) in the top cell type (rank=1) and other cell types (ranked). The pvalues were obtained using a Wilcoxon sum-rank test. Each dot represents a colocalized gene, CLU, CTSB, NR1H3 and RERE are highlighted as examples. **C)** Posterior probabilities of shared genetic signal between GWAS and eQTLs for Alzheimer’s disease, **D**) Parkinson’s disease, **E)** multiple sclerosis and **F)** schizophrenia. The closest gene to the top GWAS signal is indicated on the left, the colocalized genes is indicated on the right. The beta (Effect or OR) column indicates the effect size of the top GWAS SNP at the locus. The risk column indicates whether an increase in gene expression leads to an increase in disease risk (red) or a decrease in disease risk (blue). The LOEUF ^18^ column indicates whether the gene is constrained (low score) or not (high score).

For Alzheimer’s disease (AD) (**Figure 4C**), we found most colocalization signals in microglia (BIN1, CASS4, CD2AP, INPP5D, PICALM, RAPEB1, RIND1, SIGLEC9, TREM2, USP6NL and ZYX), which is consistent with the known pathophysiological role of microglia in AD ^30^. Several of these genes localize to the endolysosomal network (BIN1, RIN3, CD2AP, PICALM, SIGLEC9 ^31^, ZYX) ^32^, and some are known to interact (e.g. CD2AP and RIN3 ^33^ or RIN3 and BIN1 ^34^). Interestingly, two genes colocalized solely in oligodendrocytes (APH1B and CR1). APH1B is part of the γ-secretase complex, which is known to cleave APP resulting in the production of amyloid beta (Aβ), the main component of amyloid plaques (which characterize AD), while CR1 encodes the complement receptor 1, which is a regulator of complement activation. In addition, CLU, which plays a key role in Aβ clearance, aggregation and toxicity ^35^, colocalized solely in endothelial cells. We note that colocalization of a gene in a specific cell-type is not influenced by its expression levels. For example, APH1B is expressed at similar levels in all cell types (**Figure S17**) but only colocalizes with AD in oligodendrocytes, while CLU has higher expression in astrocytes than in endothelial cells but does not colocalize in astrocytes. We then leveraged allelic information from both our eQTL analysis and GWAS data to predict the effect of increasing gene expression on disease risk (**Table S5**). We found that increased expression of a number of genes was associated with an increase in AD risk (e.g. APH1B, BIN1, USP6NL) (**Figure 4C**), while an increase in expression of other genes lead to a decrease in AD risk (e.g. CASS4, PICALM or TREM2). More quantitatively, we estimated that, for example, a decrease in BIN1 expression by two standard deviations should result in an odds ratio of being diagnosed with AD of only 0.34 (**Table S5**).

Parkinson’s disease (PD) had a more complex pattern of colocalization signals (**Figure 4D**). Some genes colocalized in multiple cell types (e.g. LSM7, BIN3 or GPNMB), and some loci had colocalizations signals with multiple genes (e.g. WNT3 locus with colocalization signal for MAPT, LRRC37A2, KANSL1, ARL17B and AC0056702.2). We note that the colocalization signals at the WNT3 locus are likely due to a common inversion in the European population that is associated with PD ^36^. However, the majority of colocalized genes still colocalized in a single cell type. For example, CTSB, TOMM7 and ATP5ME colocalized only in endothelial cells. CTSB plays an essential role in lysosomal degradation of α-synuclein ^37^, while TOMM7 is a small subunit of the TOM complex that is essential for the binding of PINK1 (a gene associated with monogenic forms of the disease) to the TOM complex. For the three genes colocalized solely in endothelial cells, our results suggest that increased expression leads to a decrease in PD risk (**Figure 4D**). CD38 colocalized only in astrocytes and was shown to play an essential role in the astrocytic release of extracellular mitochondrial particles ^38^, while UBAP1 and STX4 colocalized only in oligodendrocytes. GPNMB, a gene upregulated in PD and after lysosomal stress ^39^ colocalized in both OPCs / COPs and microglia. Finally, while not reaching our colocalization threshold, we found evidence that LRRK2, a gene associated with familial forms of the disorder colocalized in microglia (PP=0.56, increased expression leading to an increase in disease risk). Our results suggest that genetic risk for PD involves a complex interaction of multiple genes acting in multiple CNS cell types.

For multiple sclerosis (MS) (**Figure 4E**), we found colocalization signals at 29 loci. AHI1, CLECL1 (recently colocalized using a large cortical eQTL data set ^17^), IQCB1, ZC2HC1A, PLEC, RPS6KA4, and TYMP colocalized in microglia. In addition, we observed colocalization signal in multiple other cell types (e.g. NR1H3 in astrocytes). As MS is primarily a disorder driven by the immune system, we caution that CNS cell type colocalization signals could be driven by pleiotropy ^40^ (i.e. a single variant or linked variants regulating different genes in different cell types). Indeed, when we tested MS genetic enrichment in cell type specific genes (derived from our control samples), we found the strongest enrichment for CNS infiltrating immune cells followed by microglia (**Figure S18**). Therefore, in order to assess the extent of pleiotropy at MS loci, we performed colocalization analysis with two immune tissues from GTEx ^2^ (blood and spleen) and 15 immune cell types from Dice ^41^ (**Figure S19**) (**Table S6**). We observed that pleiotropy between CNS cell types and immune cell types is common at MS loci. Of the 25 loci with colocalization signals in CNS cell types, we also found colocalization in immune cell types at 23 loci. The two loci that were exclusively colocalized in CNS cell types were the CD40 locus (SLC12A5 in excitatory and inhibitory neurons) and the TNFAIP8 locus (HSD17B4 in inhibitory neurons). We caution that CD40 was previously colocalized with rheumatoid arthritis at the CD40 locus in immune B cells and could be a false negative in our analysis ^42^. For the 23 loci with colocalization in both CNS cell types and immune cells, we found consistent colocalized genes at 16 loci (e.g. AHI1, CLECL1, IQCB1, PLEC, STAT4, TRAF3, TYMP, VEGFB) with, typically, additional colocalized genes in immune cell types (e.g. only IQCB1 colocalizes at the IQCB1 locus in CNS cell types, while IQCB1, EAF2 and SLC15A2 colocalize in immune cell types). In summary, we identified putative risk genes for MS in CNS cell types but caution that these results should be interpreted in light of colocalization results in immune cell types.

Schizophrenia had the most colocalization signals (**Figure 4F**) with at least one colocalized gene at 56 loci. Most colocalization signals were observed in excitatory neurons, which is consistent with the genetic enrichment of excitatory neuron specific genes in schizophrenia ^43,44^ (**Figure S18**). For 44 loci, we found a single colocalized gene, such as FURIN in excitatory neurons (previously colocalized using bulk cortical eQTL data ^45^) or CACNA1I (a voltage gated calcium channel). Other interesting colocalized genes include CUL3 (in excitatory neurons and astrocytes), which was shown to play an important role in excitation-inhibition balance ^46^, IGSF9B (in excitatory neurons), a key regulator of inhibition in the amygdala ^47^, SF3B1 (in excitatory neurons), a splicing factor subunit, which is supported by an animal model of psychosis ^48^ or TRANK1 (in excitatory neurons), which was previously shown to colocalize with bipolar disorder ^49^ and to be upregulated by valproate ^50^.

### Fine mapping of neurological disease genes risk variants to cell type regulatory elements

We next assessed whether GWAS SNPs (r^2^>0.8 with index SNP) overlapped regulatory regions ^19,20^ in close proximity of the colocalized gene (<100kb) in the colocalized cell types. We identified putative risk SNPs at more than half of the colocalized loci (56/106, **Table S7**). Notably, two AD GWAS SNPs (rs10792832 and rs3851179) overlapped a microglia specific enhancer connected to PICALM through PLAC-seq ^19^ (**Figure 5A**), while two other AD GWAS SNPs (rs10933431 and rs10933431) were located in a microglia specific enhancer connected to INPP5D through PLAC-seq (**Figure 5B**). Interestingly, INPP5D is a phosphatase which hydrolyze phosphatidylinositol-3,4,5-trisphosphate into phosphatidylinositol 3,4-diphosphate, which specifically binds to PLEKHA1 ^51^, a gene recently associated with AD through proteome-wide association study ^52^. In addition, an AD GWAS SNP (rs117618017) overlapped the APH1B promoter in oligodendrocytes (**Figure 5C**), while another (rs7515905) was located in an enhancer located within the CR1 gene body (**Figure 5D**). Similarly, we found that 2 PD GWAS SNPs (rs4698412 and rs4698413) overlapped an astrocytic enhancer located in close proximity of CD38 (**Figure 5E**), and that five PD GWAS SNPs overlapped an enhancer close to the TSS of GPNMB (4-12kb downstream). Finally, three SNPs (rs8140771, rs926334 and rs732381) overlapped a neuronal enhancer located within the CACNA1I gene body (**Figure 5G**), while five MS GWAS SNPs overlapped an astrocytic enhancer located downstream of NR1H3 (**Figure 5H**).

**Figure 5:**
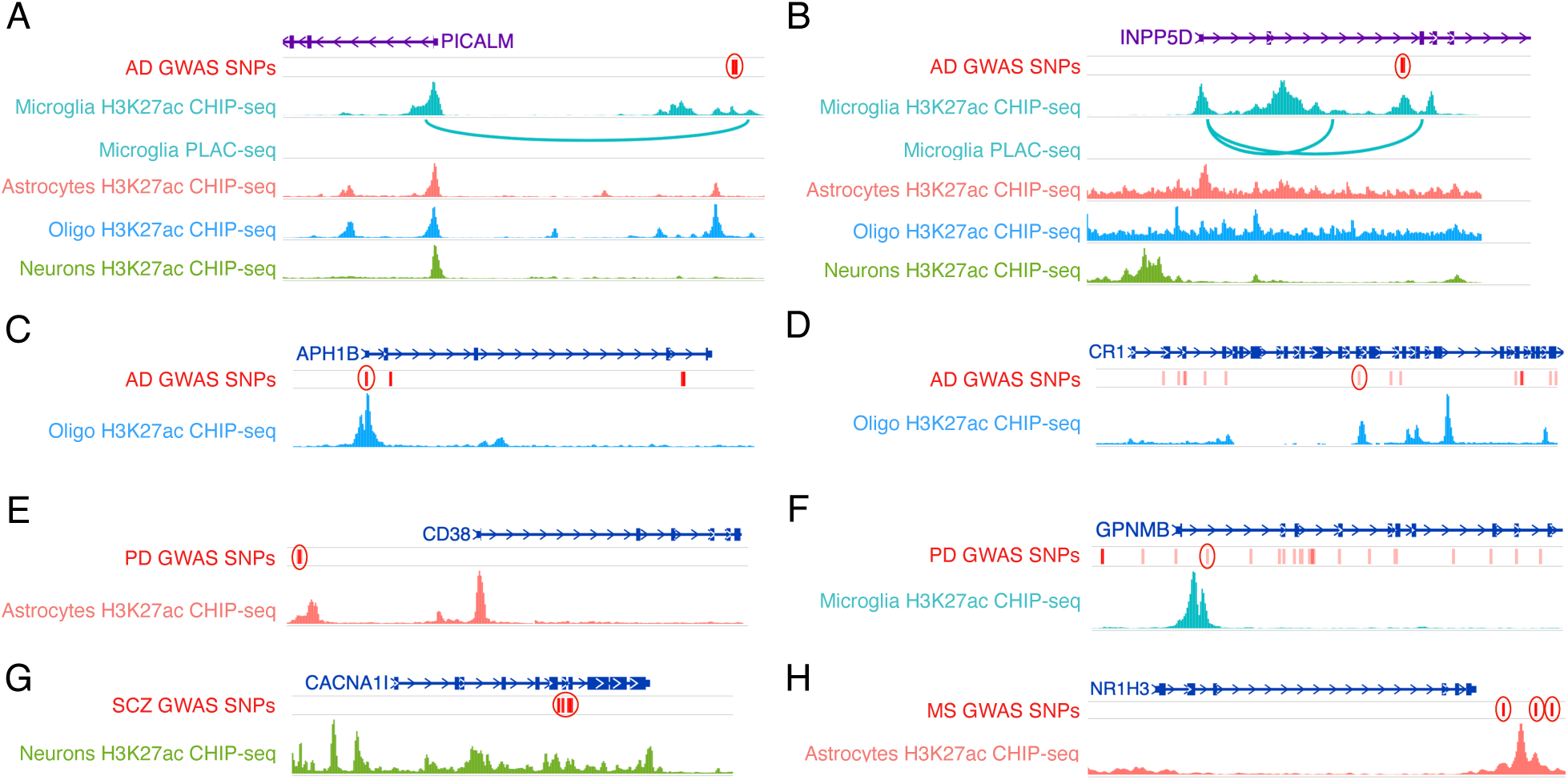
Epigenomic overlap of GWAS SNPs around colocalized genes. **A)** Genomic map indicating the location of Alzheimer GWAS SNPs (r^2^>0.8 with index SNP) overlapping a microglia specific enhancer, connected to the PICALM promoter through PLAC-seq. **B)** Genomic map indicating the location of Alzheimer GWAS SNPs (r^2^>0.8 with index SNP) overlapping a microglia specific enhancer, connected to the INPP5D promoter through PLAC-seq. **C)** Genomic map indicating the location of an Alzheimer GWAS SNP (r^2^>0.8 with index SNP) overlapping the promoter of APH1B in oligodendrocytes. **D)** Genomic map indicating the location of an Alzheimer GWAS SNP (r^2^>0.8 with index SNP) overlapping an oligodendrocyte enhancer located within the CR1 gene. **E)** Genomic map indicating the location of Parkinson’s disease GWAS SNPs (r^2^>0.8 with index SNP) overlapping an astrocyte enhancer upstream of CD38. **F)** Genomic map indicating the location of Parkinson’s disease GWAS SNPs (r^2^>0.8 with index SNP) overlapping a microglia enhancer located within GPNMB. **G)** Genomic map indicating the location of schizophrenia GWAS SNPs (r^2^>0.8 with index SNP) overlapping a neuronal enhancer located within the CACNA1I gene body. **H)** Genomic map indicating the location of multiple sclerosis GWAS SNPs (r^2^>0.8 with index SNP) overlapping an astrocytic enhancer located downstream of NR1H3.

We then used SNP2TFBS ^53^ to identify fine-mapped SNPs that could disrupt the binding of specific transcription factors (**Table S7**). Of 506 fine-mapped SNPs, we predict that 168 disrupt transcription factor binding sites. Notably, rs4663105, which is located in a microglia enhancer 24kb upstream of BIN1 (colocalized with AD), is predicted to disrupt the binding of KLF4, a transcription factor which was shown to regulate microglia activation ^54^, while rs77892763, which is located in a microglia enhancer connected to USP6NL through PLAC-seq (also colocalized with AD) is predicted to disrupt the binding of KLF5. Another example is rs2905435, the SCZ GWAS index SNP at the GATAD2A locus, which is located in the promoter region of GATAD2A, and predicted to disrupt the binding site of REST, a key transcriptional factor in neurogenesis ^55^.

In summary, our integrative analysis allowed us to fine-map disease-associated genetic variants to cell-type specific regulatory elements, highlighting potential functional mechanisms of action for disease-associated variants.

## Discussion

Here, we present the first eQTL study of all major cell types in the adult human brain by leveraging single-nucleus RNA-seq data from a large number of individuals. Integration of eQTLs with GWAS and cell-type specific regulatory elements allowed us to identify high-confidence risk genes, the cell types in which these risk genes are active, and putative genetic variants underlying risk loci for four major brain disorders. Furthermore, we leveraged allelic effects to estimate whether an increase in gene expression is associated with an increase or decrease in disease risk, an information that is crucial for drug development and often not reported.

One striking result of this study is that, at most GWAS loci, a single gene colocalized in a single cell type, allowing us to substantially refine the mechanistic hypothesis for the etiology of the different brain disorders. For example, in Alzheimer’s disease, we found most colocalization signals in microglia (e.g. BIN1 ^12,13^, PICALM ^12,13^, USP6NL ^13^), as expected. However, we also found that APH1B, a key protein from the γ-secretase complex, and CR1 colocalized only in oligodendrocytes, suggesting that oligodendrocytes play an important underrecognized role in Alzheimer’s disease etiology. Similarly, we found colocalization signals in endothelial cells only for CTSB and ATP5ME in Parkinson’s disease, or UBAP1 and STX4 in oligodendrocytes, suggesting an important role of oligodendrocytes and endothelial cells in the etiology of Parkinson’s disease.

The colocalization results appeared simplest, i.e greater number of risk genes prominently acting in few cell types, for Alzheimer’s disease compared to Parkinson’s disease, schizophrenia and multiple sclerosis, which showed more complex colocalization patterns (e.g. GPNMB locus in Parkinson’s disease, where we find colocalization with GPNMB in microglia and OPCs/COPs, NUPL2 in oligodendrocytes and TOMM7 in endothelial cells). The simplicity of Alzheimer’s colocalization patterns is consistent with the oligogenic nature of Alzheimer’s disease ^56^, and contrasts with the more polygenic architecture of schizophrenia and Parkinson’s disease ^56,57^.

For multiple sclerosis, we often found colocalization signals with different genes between CNS cell types and immune cell types, suggesting that disease-associated haplotypes regulate different genes in different cellular contexts. Future high-resolution maps of functional regulatory elements of CNS cell types from MS brain tissues might help resolve the context-specificity of genetic regulation of MS associated risk genes.

There are a number of limitations in this study: 1) the sample size is relatively small compared to eQTL studies from bulk brain tissues ^2,17^, limiting the statistical power to discover cis-eQTLs; 2) we did not measure splicing which might play an important role in disease ^58^, potentially missing a subset of colocalization signals; 3) the number of eQTL discoveries is limited for rare cell types (e.g. 23 cis-eQTL genes in pericytes), suggesting that targeted enrichments may be essential to increase the number of cis-eQTLs discoveries in rare cell populations; and 4) the GWAS used were all related to disease risk and not disease progression. Hence, colocalized genes might not be therapeutically relevant at the time of diagnosis.

In summary, our study provides a systematic investigation of eQTLs in cell-types of the adult human brain, defines a reference data set of CNS cell-type specific eQTLs and provides a foundational resource of high-confidence colocalized genes in disease-relevant cell types for robust future functional studies of neurodegenerative disease mechanisms using appropriate iPSC-based human cell models.

## Data Availability

A shinyApp to browse the result of this study is available at: https://malhotralab.shinyapps.io/brain_cell_type_eqtl/

The full eQTL summary statistics are available on zenodo at: https://doi.org/10.5281/zenodo.5543734

All single nuclei RNA-seq and genotypes will be made available after peer-review.

https://malhotralab.shinyapps.io/brain_cell_type_eqtl/

https://doi.org/10.5281/zenodo.5543734

## Online Methods

### Nuclei isolation and single nuclei RNA sequencing

We generated brain single nuclei RNA-seq data for two datasets (Roche_MS, Roche_AD, described below), that we complemented with raw sequencing data from three published datasets (Columbia_AD ^16^, Mathys_AD ^14^ and Zhou_AD ^15^) (**Table S1**). Samples originated from the prefrontal cortex (Roche_MS, Columbia_AD ^16^, Mathys_AD ^14^ and Zhou_AD ^15^), temporal cortex (Roche_AD) and deep white matter (Roche_MS, Roche_AD and Columbia_AD). A description of the nuclei isolation and single nuclei RNA-sequencing for the unpublished datasets is provided below:

The Roche multiple sclerosis dataset (Roche_MS) consists of 166 samples from 83 unique individuals (29 controls and 54 cases). For cases, samples were taken from multiple different lesion types (if available) assessed by a neuropathologist (normal appearing white matter (NAWM), active lesions (AL), chronic active lesions (CAL), chronic inactive lesions (CIL), remyelinating lesions (RL), normal appearing grey matter (NAGM), grey matter lesions (GML)).

The Roche Alzheimer’s disease dataset (Roche_AD) consists of 80 samples from 40 unique individuals (one sample from the temporal cortex and one from deep white matter for each individual).

For both datasets, nuclei were isolated from fresh-frozen 10μm sections, using Nuclei Pure Prep Nuclei Isolation Kit (Sigma Aldrich) with the following modifications. The regions of interest were macro-dissected with a scalpel blade, lysed in Nuclei Pure Lysis Solution with 0.1% Triton X, 1mM DTT and 0.4U/ul SUPERase-In(tm) RNase Inhibitor (ThermoFisher Scientific) freshly added before use, and homogenized with the help first of a 23G and then of a 29G syringe. Cold 1.8M Sucrose Cushion Solution, prepared immediately before use with the addition of 1mM DTTand 0.4U/ul SUPERase-In(tm) RNase Inhibitor, was added to the suspensions before they were filtered through a 30μm strainer. The lysates were then carefully and slowly layered on top of 1.8M Sucrose Cushion Solution previously added in new Eppendorf tubes. Samples were centrifuged for 45 minutes at 16000xg at 4°C. Pellets were re-suspended in Nuclei Storage Buffer with 0.4U/ul SUPERase-In(tm) RNase Inhibitor, transferred in new Eppendorf tubes and centrifuged for 5 minutes at 500xg at 4°C. Pellets were again re-suspended in Nuclei Storage Buffer with 0.4U/ul SUPERase-In(tm) RNase Inhibitor, and centrifuged for 5 minutes at 500xg at 4°C. Finally, purified nuclei were re-suspended in Nuclei Storage Buffer with 0.4U/ul SUPERase-In(tm) RNase Inhibitor, stained with trypan blue and counted using Countess II (Life technology). A total of 12,000 estimated cells from each sample were loaded on the 10x Single Cell Next GEM G Chip. cDNA libraries were prepared using the Chromium Single Cell 3’ Library and Gel Bead v3 kit according to the manufacturer’s instructions. cDNA libraries were sequenced using the Illumina NovaSeq 6000 System and NovaSeq 6000 S2 Reagent Kit v1.5 (100 cycles), aiming at a sequencing depth of minimum 30K reads/nucleus.

### Single nuclei RNA-seq analysis

All samples from the Roche_MS datasets were processed with CellRanger (v3.1.0), using the GRCh38 reference human genome and the ensembl Homo_sapiens GRCh38.96 reference annotation (modified to count intronic reads). Gene expression quantifications for each nucleus were obtained from the ‘filtered_feature_bc_matrix’ CellRanger (v3.1.0) output folder. We identified doublets using scDblFinder ^59^ (version 1.4.0), applied to each sample separately. After removing doublets, we also removed nuclei with less than 300 features and 500 counts, as well as samples with less than 500 cells. We did quality control with SampleQC with default parameters (version 0.4.5) ^60^. SampleQC allowed us to identify a subset of cells in many samples with both high splice ratios and high mitochondrial proportions (90% of reads being spliced), which were excluded. After excluding outliers, we again excluded any samples with fewer than 500 cells remaining, resulting in 750,614 nuclei across 166 samples passing QC. Samples were integrated using conos ^61^ (50 PCs, resolution=8) and broad clusters were annotated based on the expression of canonical markers (SLC17A7 for excitatory neurons, GAD2 for inhibitory neurons, AQP4 for astrocytes, MOG for oligodendrocytes, PDGFRA for oligodendrocytes precursors cells and committed oligodendrocytes precursors (OPCs / COPs), C1QA for Microglia, CLDN5 for endothelial cells and RGS5 for Pericytes).

All samples from the AD datasets (Roche_AD, Columbia_AD, Mathys_AD ^14^ and Zhou_AD ^15^) were processed with CellRanger (v3.1.0), using the GRCh38 reference human genome and the ensembl Homo_sapiens GRCh38.91 reference annotation (modified to count intronic reads). Nuclei were defined as barcodes with at least 500 unique molecular identifiers (UMI) (excluding mitochondrial RNA) and less than 5% of mitochondrial RNA. If a sample had more than 10k nuclei, we kept the 10k nuclei with the highest number of UMI. Doublet were identified using scDblFinder ^59^ (version 1.4.0). Data from the different samples were integrated using both Conos ^61^ (50 PCs, resolution=8) and Harmony ^62^ (30 PCs, resolution=0.8) and labelled independently using the canonical markers described above. A subset of small clusters from the Conos integration expressed multiple canonical markers and were labeled as “mixed”. Nuclei that were not labeled as “mixed” were annotated with the Conos cell type labels. The remaining nuclei were labeled with the Harmony cell type label.

Pseudo-bulk gene expression matrices were then generated by summing all counts for each gene in each patient in each cell type and normalized by scaling the total counts per patient for each cell type to 1 million.

### Genotyping QC

We genotyped samples from the Roche_AD and Roche_MS datasets using the GSAv3 illumina CHIP. Genotypes were then imputed using the Haplotype Reference Consortium (HRC) reference panel (version r1.1) ^63^ and lifted over to GRCh38. Genotype processing and quality control was performed using Plink v1.9^56^. SNPs with imputation score <0.4 or with missingness greater than 5% were excluded, as well as individuals with more than 2% of missing genotypes. In addition, we obtained whole genome sequencing results for the ROSMAP datasets (Columbia_AD ^16^, Mathys_AD ^14^, Zhou_AD ^15^) ^64^. Similarly, SNPs with missingness greater than 5% were excluded, as well as individuals with more than 2% of missing genotypes. Genetic variants that were common to imputed genotypes and whole genome sequencing were then merged. Post-merging, SNPs with minor allele frequency (MAF)<5% or deviating from Hardy-Weinberg equilibrium (Pvalue <1e-6) were excluded. We identified related individuals (pi_hat>0.2) and only kept one individual from related pairs. In addition, individuals deviating from more than three standard deviations from 1000 genomes European populations^57^ on the first and second components of a multidimensional scaling (MDS) reduction were excluded (**Figure S20**). Finally, only individuals with both genotype and single nuclei RNA-seq were retained for the eQTL analysis. After QC, we obtained high quality genotypes for ~5.3 million SNPs (MAF>5%) in 196 individuals.

### eQTL mapping

We mapped cis-eQTL within a 1 megabase (MB) window of the TSS of each gene expressed in at least 5% of the nuclei (belonging to a broad cell type) using fastQTL ^65^ (7208-10846 genes across the 8 cell types). We used the following covariates: 3 first genotyping principal components (PCs), disease status (MS, AD, control or other), study (Roche_MS, Roche_AD, Columbia_AD ^16^, Mathys_AD ^14^ and Zhou_AD ^15^) and the 90 first expression PCs. This number of expression PCs maximized the number of cis-eQTL detected (**Figure S21**). While the number of expression PCs is high in comparison to bulk RNA-seq eQTL studies, the results appear robust for multiple reasons: 1) eQTL strongly replicated in the metabrain ^17^ bulk RNA-seq eQTL study (**Figure 2C and S3A**), 2) were enriched close to the TSS (**Figure 2D**), 3) affected less constrained genes (**Figure 2D**), 4) were enriched around cell type specific epigenomic marks (**Figure 3A**), 5) the pvalue distribution were sensible (**Figure S2**), and, 6), random permutation of the gene expression labels lead to only two genes with a significant eQTL (5% FDR). Multiple testing correction was performed using the qvalue R package ^25^ on the gene-level pvalues (bpval). Estimates of the proportion of true alternative hypothesis (i.e. proportion of genes with a cis-eQTL, **Figure S2**) was performed using the pi1 statistic from the qvalue R package ^25^. Independent cis-eQTL mapping was done using QTLtools ^66^, with the same covariates and window size as the original cis-eQTL analysis.

### Sharing of brain cell type eQTL with metabrain cortex eQTL

We obtained p-values from cortical samples (N=2970) from the metabrain study ^17^ corresponding to the most significant SNP-gene pairs for each cis-eQTL gene (N=6108 at 5%FDR). Sharing of eQTL was computed using the pi1 statistic from the qvalue R package ^25^ or as the proportion of metabrain SNP-gene pairs with a p-value <0.05.

### Epigenome enrichments

We used QTLtools ^66^(fdensity) to test whether cell type specific regulatory elements were enriched around our cell type cis-eQTL (N=6108). For each 10kb bin in a 2MB window around the cis-eQTL, fdensity computes the number of functional elements overlapping the bin. The epigenomic data were obtained from three different studies (Nott et al. ^19^, Corces et al. ^20^ and the DLPFC region for Fullard et al. ^23^), and consists of ATAC-seq (bulk ^23^ or single nuclei ^20^) and CHIP-seq ^19^(H3K4me3 and H3K27ac) from diverse brain cell types. For each dataset, we defined cell type specific epigenomic marks as epigenomic marks observed in a single cell type. Epigenomic regions were lifted over to hg38 (if not provided for this genome build by the authors). We repeated the enrichment analysis (fdensity) for neuronal cis-eQTL using different filters. None of the filters (listed below) showed an enrichment of neuron specific regulatory elements around neuron cis-eQTL: 1) Restricting to most significant eQTL in neurons (0.1% FDR), 2) eQTL detected in neurons with 20 PCs (instead of 90) (5%FDR), 3) eQTLs for genes expressed in at least 20% of the nuclei (instead of 5%), 4) eQTLs only significant in neurons (5% FDR), 5) cell type specific eQTLs using our negative binomial model (5% FDR), and 6) eQTLs obtained using stringent QC criteria at nuclei level (at least 1200 UMI or at least 1500 UMI).

### Interaction model

We tested whether the effect size of our cis-eQTL (N=6108, 5% FDR) were cell type specific using a negative binomial mixed model (as implemented in the R package glmmTMB ^67^). First, we tested whether the effect sizes of the eQTL in the detected cell types were different from the average effect sizes in all other cell types. For each gene, the model used was the following: raw counts ~ genotype (0,1,2) + cell_type + genotype:cell_type_eqtl (cell type in which the eQTL was discovered, coded as 0,1) + PC 1-3 genotype matrix + PC 1-5 expression matrix + study + disease_status + (1|individual) (random effect), using as offset the log of the library size. The pvalue of the interaction effect was then corrected for multiple testing using the Benjamini and Hochberg procedure^61^. Second, we tested whether the effect sizes of the eQTL in the detected cell types were different from at least one other cell type. We used a similar model as described above, except that the specified interaction term was “genotype*cell_type” instead of “genotype:cell_type_eqtl”. Interaction pvalues were Bonferroni corrected across cell types (for each gene) and the minimum corrected pvalue was retained. These corrected pvalues were then further corrected for multiple testing using the Benjamini and Hochberg procedure ^68^. Finally, we used the same results but retained the maximum pvalue of the interaction terms (for each gene) in order to assess whether the cis-eQTL in the discovered cell type had a different effect than all other cell types. These pvalues were then corrected for multiple testing with the Benjamini and Hochberg procedure ^68^.

### Fine-mapping of cis-eQTLs

We used CaveMan ^69^ to fine-map our cis-eQTLs. Briefly, CaveMan performs 10,000 eQTL analysis after bootstrapping the data (i.e. new datasets are created with randomly picked individuals with replacement). CaveMan then records the proportion of times that a given SNP is ranked among the top 10 most associated SNPs. These proportions are used to compute the CaVEMaN score, which is then calibrated to estimate the causal probability for each SNP.

### GWAS summary statistics

We obtained publicly available GWAS summary statistics for Alzheimer’s disease ^4^, multiple sclerosis ^29^ and schizophrenia ^28^. For Parkinson’s disease, we performed an inverse-variance-weighted meta-analysis ^70^ using summary statistics from Nalls et al. 2014 ^26^ and Nalls et al. 2019 ^27^.

### Heritability enrichment

We used the normalized (CPM) pseudo-bulk gene expression matrix from control individuals from our MS_Roche dataset for this analysis (N=83). For each gene, we computed the average CPM across samples resulting in one expression value for each gene in each cell type. We only retained genes with a mean CPM value greater than 1 and genes for which we had a gene-level genetic enrichment estimate from MAGMA ^71^. We then computed the proportion of the total expression of each gene across the different cell types (i.e. mean_cpm/sum(mean_cpm)). This captures how specific a gene is to a given cell type (e.g. 92% of AQP4 expression was observed in astrocytes). Finally, we tested the top 1000 most specific genes (for each cell type) for enrichment in GWAS associations using MAGMA ^71^.

### Colocalization

For the Alzheimer ^4^, Parkinson ^26,27^ and multiple sclerosis ^29^ GWAS, we defined loci as the coordinates of the most extreme coordinates of SNPs in LD (r^2^>0.1 in 1000 genomes ^72^ EUR individual) with the reported index SNPs using LDlinkR ^73^. For the schizophrenia GWAS ^28^, we used the reported loci coordinates (r^2^>0.1 with index SNP). For each loci, we then tested colocalization between the GWAS and eQTL signals for genes with at least 10 SNPs using the “coloc.abf” function of the Coloc R package^8^ (with default prior). Minor allele frequencies were derived from our eQTL data set for colocalization analysis with the CNS eQTLs. For GTEx^7^, we used the reported MAF from the GTEx consortium. For DICE ^41^, we used the MAF of the 1000 genomes European reference population ^72^. Allelic directions and estimates of the effect of increasing gene expression by one standard deviation on disease risk were derived from the effect sizes of the SNP with the most significant eQTL pvalue (and most significant GWAS pvalue for ties).

### Transcription factor SNP-motif association

SNP2TFBS ^53^ is a resource indicating whether common genetic variants disrupt transcription factor binding sites. We used the webtool available at: https://ccg.epfl.ch/snp2tfbs/snpselect.php to test whether our list of GWAS linked SNPs (r^2^>0.8 with index SNP) overlapping an epigenomic mark in close proximity to a colocalized gene (<100kb) could potentially disrupt a transcription factor binding site. Briefly, SNP2TFBS scores motifs from transcription factors (using position weight matrices) for both alleles at each SNP position and outputs whether SNPs are predicted to abolish, create or change the affinity of one or several transcription factor (TF) binding sites.

## Data availability

The full eQTL summary statistics are available on zenodo at: https://doi.org/10.5281/zenodo.5543734

## Acknowledgement

The results published here are in whole or in part based on data obtained from the AD Knowledge Portal (https://adknowledgeportal.org). Study data were provided by the Rush Alzheimer’s Disease Center, Rush University Medical Center, Chicago. Data collection was supported through funding by NIA grants P30AG10161 (ROS), R01AG15819 (ROSMAP; genomics and RNAseq), R01AG17917 (MAP), R01AG30146, R01AG36042 (5hC methylation, ATACseq), RC2AG036547 (H3K9Ac), R01AG36836 (RNAseq), R01AG48015 (monocyte RNAseq) RF1AG57473 (single nucleus RNAseq), U01AG32984 (genomic and whole exome sequencing), U01AG46152 (ROSMAP AMP-AD, targeted proteomics), U01AG46161(TMT proteomics), U01AG61356 (whole genome sequencing, targeted proteomics, ROSMAP AMP-AD), the Illinois Department of Public Health (ROSMAP), and the Translational Genomics Research Institute (genomic). Additional phenotypic data can be requested at www.radc.rush.edu.

## Author contributions

JB and DM designed the study and wrote the manuscript. JB mapped raw sequencing data, performed QC on the AD datasets and performed the eQTL, coloc and fine-mapping analysis. DC generated single-nucleus RNAseq data on AD and MS samples. WM performed the quality control on the MS dataset and provided critical statistical comments. LF provided Alzheimer’s samples. AW, EU and SA provided MS samples. WO, VAI and SS genotyped MS and a subset of AD samples and provided imputed genotype data. VM and PDJ provided snucRNAseq and whole genome sequencing data on a subset of AD samples and provided critical inputs on the interpretation of the results.

## Supplementary figures legends

**Figure S1**: Mean expression across sample

The mean count per million (CPM) per cluster are shown for canonical markers of cell type identity (z-scaled for each cell type).

**Figure S2**: Distribution of cis-eQTL pvalues

Histograms of the cis-eQTL pvalues for each gene for the different cell types. The pi1 statistic estimates the proportion of true alternative hypothesis (i.e. the proportion of genes with a cis-eQTL).

**Figure S3**: Replication of cis-eQTLs in a large cortical eQTL study

**A)** Histograms of cortical SNP-gene pvalues matching our cis-eQTLs. The ‘prop’ label indicates the proportion of pvalues that are below 0.05, while the ‘pi1’ label indicates an estimate of the proportion of cis-eQTLs that replicate. **B**) LOEUF score (a measure of whether genes are constrained) for replicating cis-eQTLs (p<0.05) and non-replicating cis-eQTLs (p>0.05). Lower scores indicate that genes are more constrained. **C**) Absolute distance to the TSS for replicating (p<0.05) and non-replicating cis-eQTLs (p>0.05).

**Figure S4**: Location of cis-eQTLs

The number of cis-eQTLs that are located upstream, downstream or within the gene body of the affected gene are shown.

**Figure S5**: Rank of affected cis-eQTL genes.

The proportion of cis-eQTLs affecting the closest gene (rank =1), second closest gene (rank=2), third closest gene (rank=3), fourth closest gene (rank=4), fifth closest gene (rank=5), or further (rank>5) are shown for each cell type.

**Figure S6**: Expression level of cis-eQTL genes.

**A**) Median CPM (log2 +1) are shown for genes with a cis-eQTL and genes without a cis-eQTLs (for all glial or neuronal cell types. **B**) Median CPM (log2 +1) are shown for genes with a cis-eQTL and genes without a cis-eQTLs (for all cell types). Pvalues were obtained using a Wilcoxon rank-sum test and are not corrected for multiple testing.

**Figure S7**: Independent cis-eQTL discoveries and properties.

**A)** Schematic representation of the effect of an additional independent SNP on gene expression.

**B)** Number of independent cis-eQTLs (5% FDR) for each cell type. **C**) Distribution of independent cis-eQTLs around the TSS. **D**) Absolute distance to the TSS for the main cis-eQTL and the secondary cis-eQTL.

**Figure S8**: Fine-mapping of cis-eQTLs.

**A)** Proportion of fine-mapped SNPs overlapping and epigenomic mark for different causal probability thresholds. **B**) Distribution of causal probabilities for cis-eQTLs in the different cell types.

**Figure S9**: Enrichment of cell type specific epigenomic marks around cis-eQTLs (Nott) Enrichment of cell type specific epigenomic marks (CHIP-seq of H3K4me3 and H3K27ac) around cis-eQTLs for each cell type. The number of cell type specific epigenomic marks was computed for each 10kb around the cis-eQTLs in a 2MB window.

**Figure S10**: Enrichment of cell type specific epigenomic marks around cis-eQTLs (Fullard) Enrichment of cell type specific epigenomic marks (ATAC-seq on NeuN+ and NeuN-sorted nuclei) around cis-eQTLs for each cell type. The number of cell type specific epigenomic marks was computed for each 10kb around the cis-eQTLs in a 2MB window.

**Figure S11**: Absolute distance to the TSS for genes with a cell type specific genetic effect

**A**) Absolute distance to the TSS for cis-eQTL genes with (and without) a different genetic effect in the discovered cell type than the aggregate genetic effect in all other cell types (5%FDR). **B**) Absolute distance to the TSS for cis-eQTL genes with (and without) a different genetic effect in the discovered cell type than at least one other cell type (5%FDR). **C**) Absolute distance to the TSS for cis-eQTL genes with (and without) a different genetic effect in the discovered cell type than all other cell types (5%FDR).

**Figure S12**: Constraint scores for cell type specific cis-eQTLs

**A**) LOEUF scores for genes with (and without) a cis-eQTL with a different genetic effect in the discovered cell type than the aggregate genetic effect in all other cell types (5%FDR). **B**) LOEUF scores for genes with and without a cis-eQTL with a different genetic effect in the discovered cell type than at least one other cell type (5%FDR). **C**) LOEUF scores for genes with and without a cis-eQTL with a different genetic effect in the discovered cell type than all other cell types (5%FDR).

**Figure S13**: Genetic enrichment of cell type specific cis-eQTLs

Genetic enrichment pvalue obtained using MAGMA for genes with a cis-eQTLs that have a different effect size in the discovered cell types than all other cell types (192 genes in total at 5% FDR).

**Figure S14**: Examples of cell type specific cis-eQTLs.

The level of expression for different genotypes is shown for four different genes. Each dot represents one individual. The displayed pvalue is the interaction pvalue testing whether the genetic effect in the discovered cell type is different than the aggregate genetic effects across all other cell types.

**Figure S15**: Distribution of interaction pvalues for cell type specific cis-eQTLs

Histograms of interaction pvalues testing for cell type specific effects. For cis-eQTLs (5% FDR) discovered in each cell type (rows), we tested whether the genetic effect for the same SNP-gene were significantly different in the other cell types (columns). The pi1 statistic^25^ estimated the proportion of true alternative hypothesis (i.e. the proportion of SNP-gene pairs with a different genetic effect in the tested cell type).

**Figure S16**: Number of colocalized loci

Number of colocalized loci (blue, posterior probability >0.7) and total number of loci tested (red).

**Figure S17**: Expression of Alzheimer colocalized genes

Median log2(CPM+1) for each colocalized genes in each cell type across the 196 individuals.

**Figure S18**: Genetic enrichment of cell type specific genes

Enrichment strength (-log10P) of the top 1000 most specific genes in each cell type in genetic associations from the GWAS of the different traits.

**Figure S19**: Colocalization results for multiple sclerosis in immune tissues and cell types Posterior probabilities of shared genetic signal between the multiple sclerosis GWAS and eQTL discovered in immune tissues from GTEx^7^ (blood, spleen) and immune cell types. The OR column indicates the odds ratio of the top GWAS SNP at the locus. The closest gene to the top GWAS signal is indicated on the left, the colocalized genes is indicated on the right. The risk column indicates whether an increase in gene expression leads to an increase in disease risk (red), a decrease in disease risk (blue) or whether the colocalization signal is due to a splicing QTL (orange). The LOEUF column indicates whether the gene is constrained (low score) or not (high score).

**Figure S20**: Multidimensional scaling of genotyping data

MDS1 and MDS2 of our genotyped samples (in pink) with individuals from the 1000 genomes project. The red squares indicate 3 standard deviations on MDS1 and MDS2 derived from EUR individuals from the 1000 genomes project.

**Figure S21**: Number of cis-eQTLs discovered in function of number of principal components Number of cis-eQTLs discovered (5% FDR) in function of the number of gene expression principal components used as covariate for the 8 major brain cell types.

## Supplementary tables legends

**Table S1**: Number of samples/individuals per dataset

Number of samples and individuals from the 5 different datasets we used in this study.

**Table S2**: cis-eQTLs results

Cis-eQTL analysis results for all genes tested in 8 major brain cell types. Only the best SNP per gene is reported (even if not significant).

**Table S3**: Fine-mapping results

Fine-mapping results for cis-eQTL genes (5% FDR).

**Table S4**: Cell type specific cis-eQTLs results

Results from our interaction models testing for a cell type specific genetic effect.

**Table S5**: Colocalization results

Colocalization results for Alzheimer’s disease, Parkinson’s disease, schizophrenia and multiple sclerosis in cell types from the central nervous system.

**Table S6**: Colocalization results for multiple sclerosis in immune tissues/cell types Colocalization results for multiple sclerosis in GTEx immune tissues (blood and spleen) and immune cell types from Dice.

**Table S7**: Epigenomic overlap of GWAS SNPS around colocalized genes

Epigenomic marks overlapping a GWAS SNP (r^2^>0.8 with index SNP) within 100kb from a colocalized gene in the colocalized cell type for Alzheimer’s disease, Parkinson’s disease, schizophrenia and multiple sclerosis.

