## Supplementary figures for "Cell-type specific cis-eQTLs in eight brain cell-types identifies novel risk genes for human brain disorders"

### Figure S1

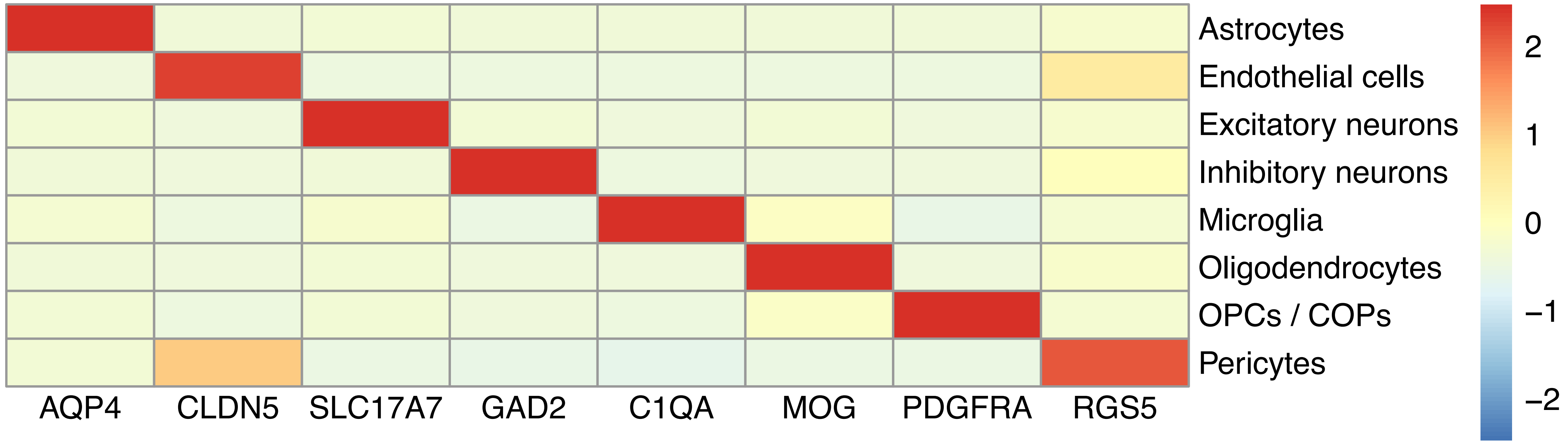
Astrocytes
Endothelial cells
Excitatory neurons
Inhibitory neurons
Microglia
Oligodendrocytes
OPCs / COPs
Pericytes
AQP4
CLDN5
SLC17A7
GAD2
C1QA
MOG
PDGFRA
RGS5
2
1
0
−1
−2

### Figure S2

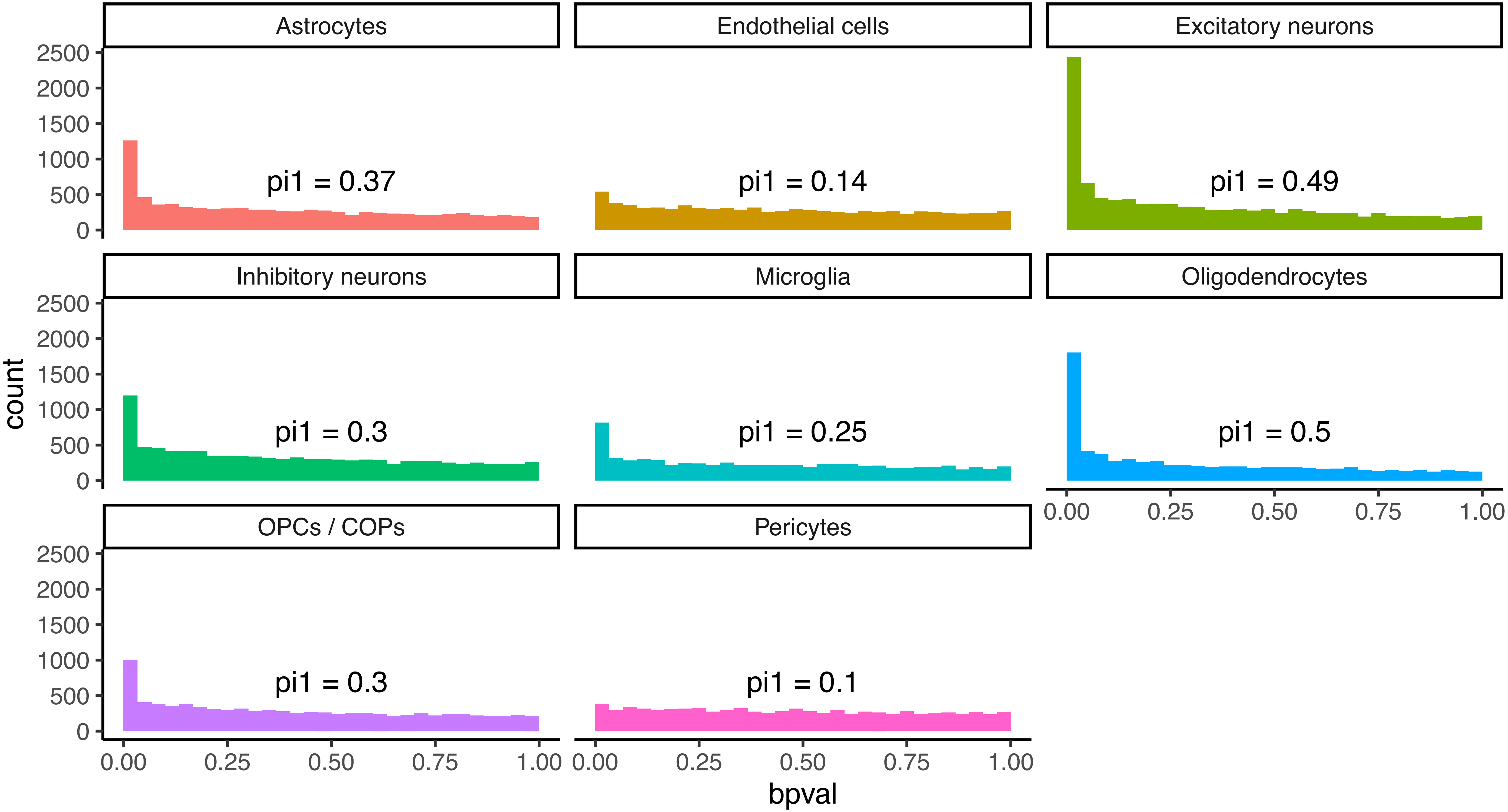

Astrocytes
Endothelial cells
Excitatory neurons
Inhibitory neurons
Microglia
Oligodendrocytes
OPCs / COPs
Pericytes
pi1 = 0.37
pi1 = 0.14
pi1 = 0.49
pi1 = 0.3
pi1 = 0.25
pi1 = 0.5
pi1 = 0.3
pi1 = 0.1
count
0
500
1000
1500
2000
2500
0.00
0.25
0.50
0.75
1.00
bpval

### Figure S3

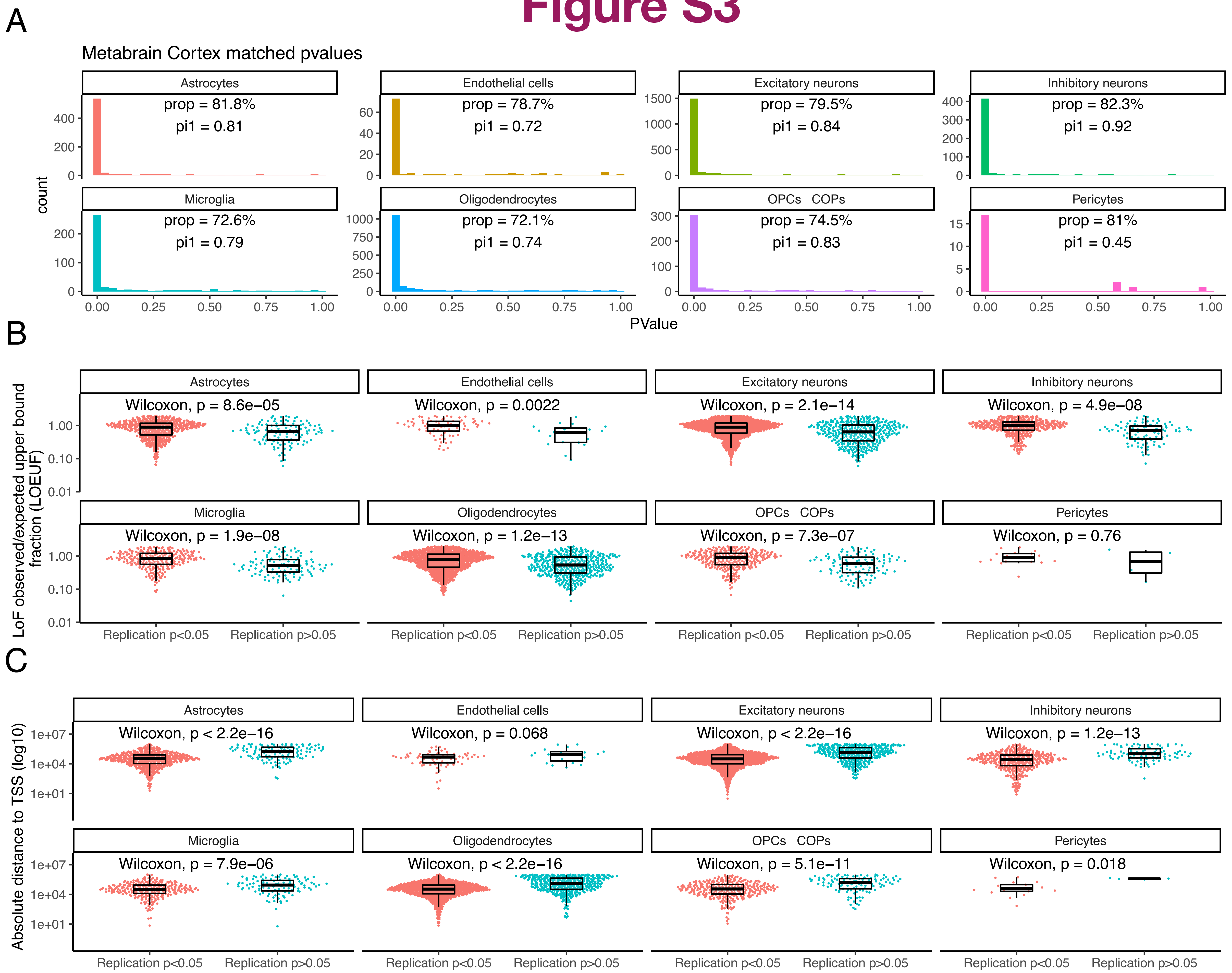

A
Metabrain Cortex matched pvalues
count
PValue
Astrocytes
prop = 81.8%
pi1 = 0.81
Endothelial cells
prop = 78.7%
pi1 = 0.72
Excitatory neurons
prop = 79.5%
pi1 = 0.84
Inhibitory neurons
prop = 82.3%
pi1 = 0.92
Microglia
prop = 72.6%
pi1 = 0.79
Oligodendrocytes
prop = 72.1%
pi1 = 0.74
OPCs COPs
prop = 74.5%
pi1 = 0.83
Pericytes
prop = 81%
pi1 = 0.45
B
LoF observed/expected upper bound fraction (LOEUF)
Astrocytes
Wilcoxon, p = 8.6e−05
Endothelial cells
Wilcoxon, p = 0.0022
Excitatory neurons
Wilcoxon, p = 2.1e−14
Inhibitory neurons
Wilcoxon, p = 4.9e−08
Microglia
Wilcoxon, p = 1.9e−08
Oligodendrocytes
Wilcoxon, p = 1.2e−13
OPCs COPs
Wilcoxon, p = 7.3e−07
Pericytes
Wilcoxon, p = 0.76
Replication p<0.05
Replication p>0.05
C
Absolute distance to TSS (log10)
Astrocytes
Wilcoxon, p < 2.2e−16
Endothelial cells
Wilcoxon, p = 0.068
Excitatory neurons
Wilcoxon, p < 2.2e−16
Inhibitory neurons
Wilcoxon, p = 1.2e−13
Microglia
Wilcoxon, p = 7.9e−06
Oligodendrocytes
Wilcoxon, p < 2.2e−16
OPCs COPs
Wilcoxon, p = 5.1e−11
Pericytes
Wilcoxon, p = 0.018
Replication p<0.05
Replication p>0.05

### Figure S4

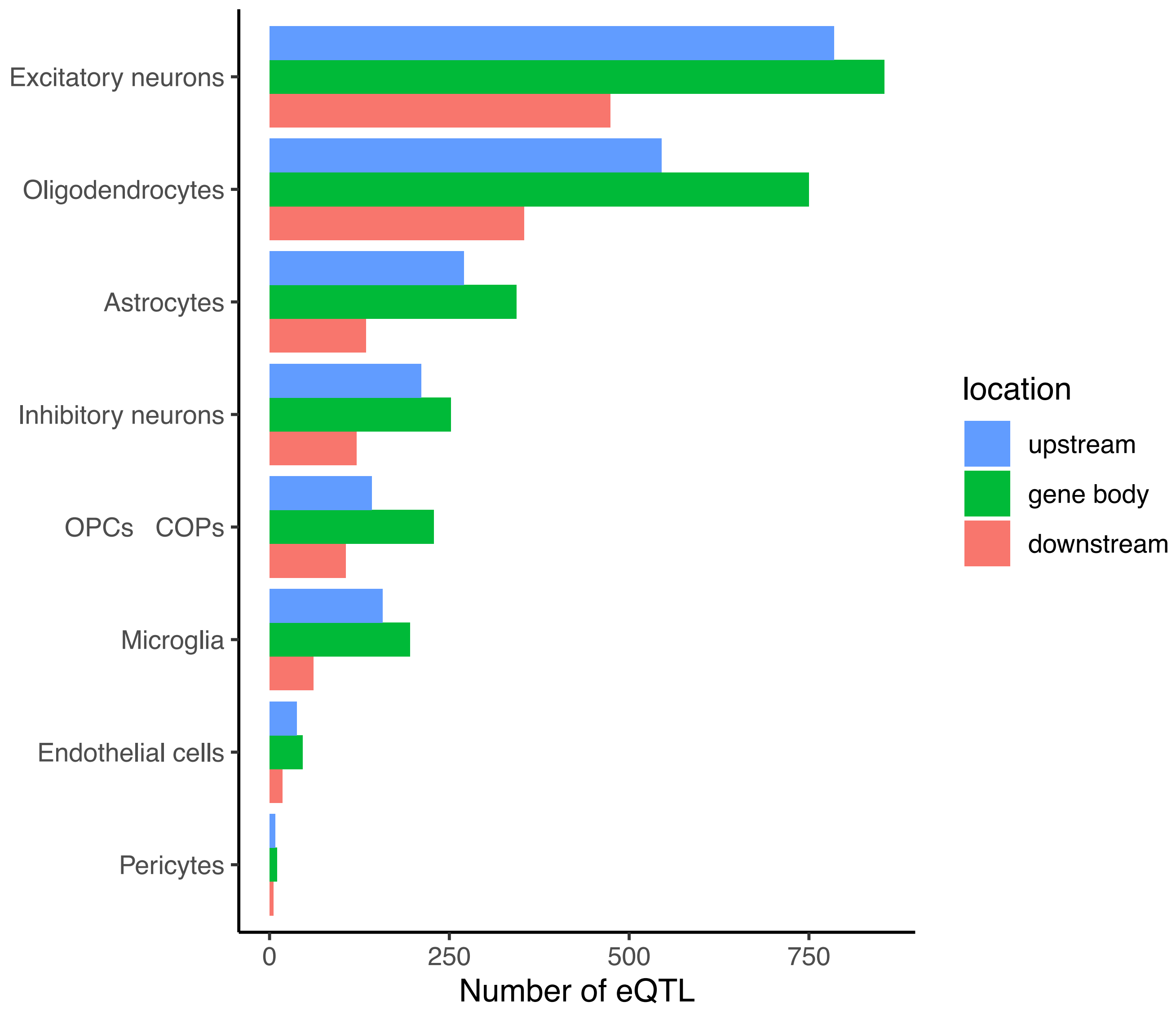
Excitatory neurons
Oligodendrocytes
Astrocytes
Inhibitory neurons
OPCs COPs
Microglia
Endothelial cells
Pericytes
0
250
500
750
Number of eQTL
location
upstream
gene body
downstream

### Figure S5

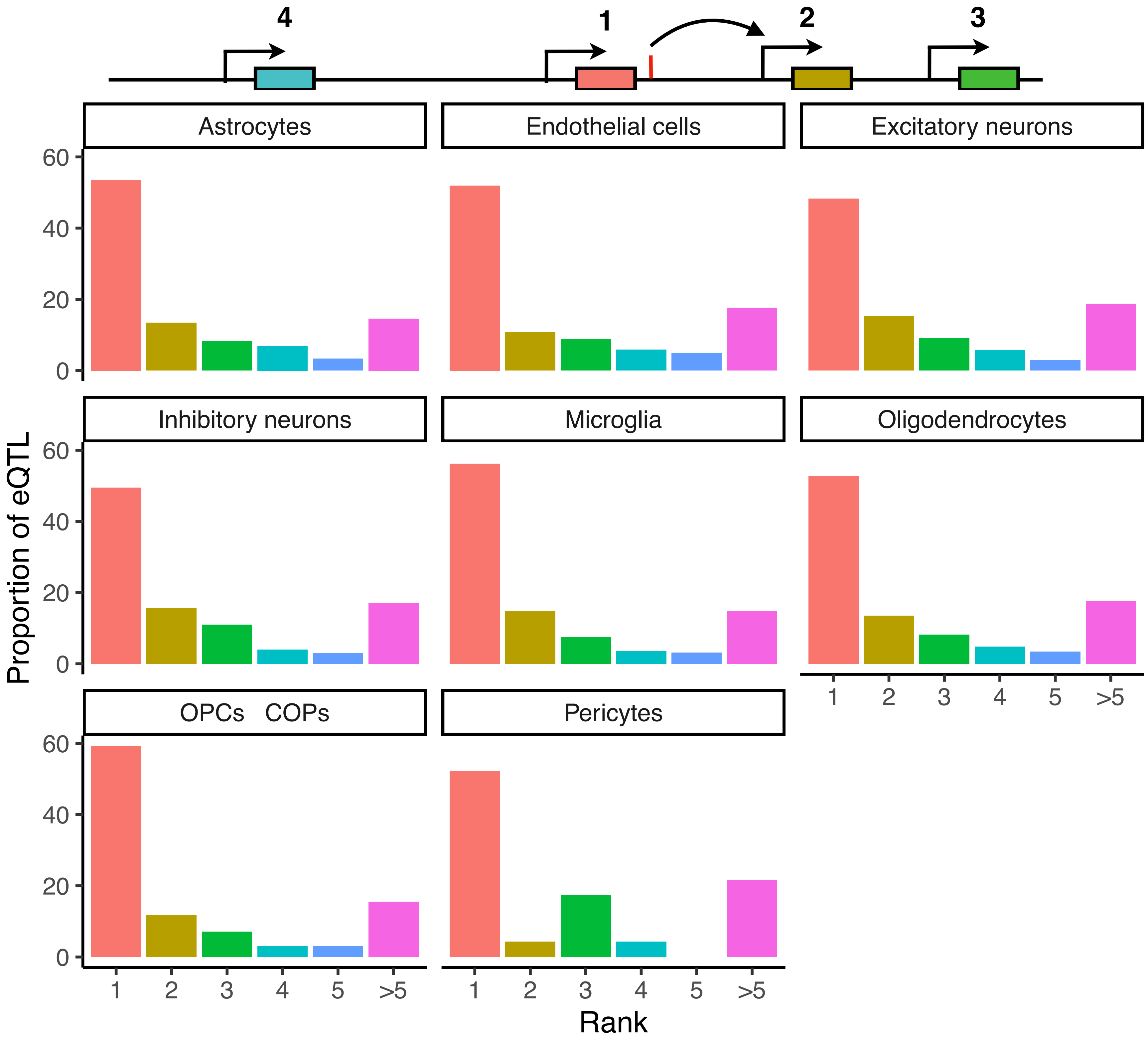

4
1
2
3
Astrocytes
Endothelial cells
Excitatory neurons
Inhibitory neurons
Microglia
Oligodendrocytes
OPCs COPs
Pericytes
Proportion of eQTL
60
40
20
0
1
2
3
4
5
>5
Rank

### Figure S6

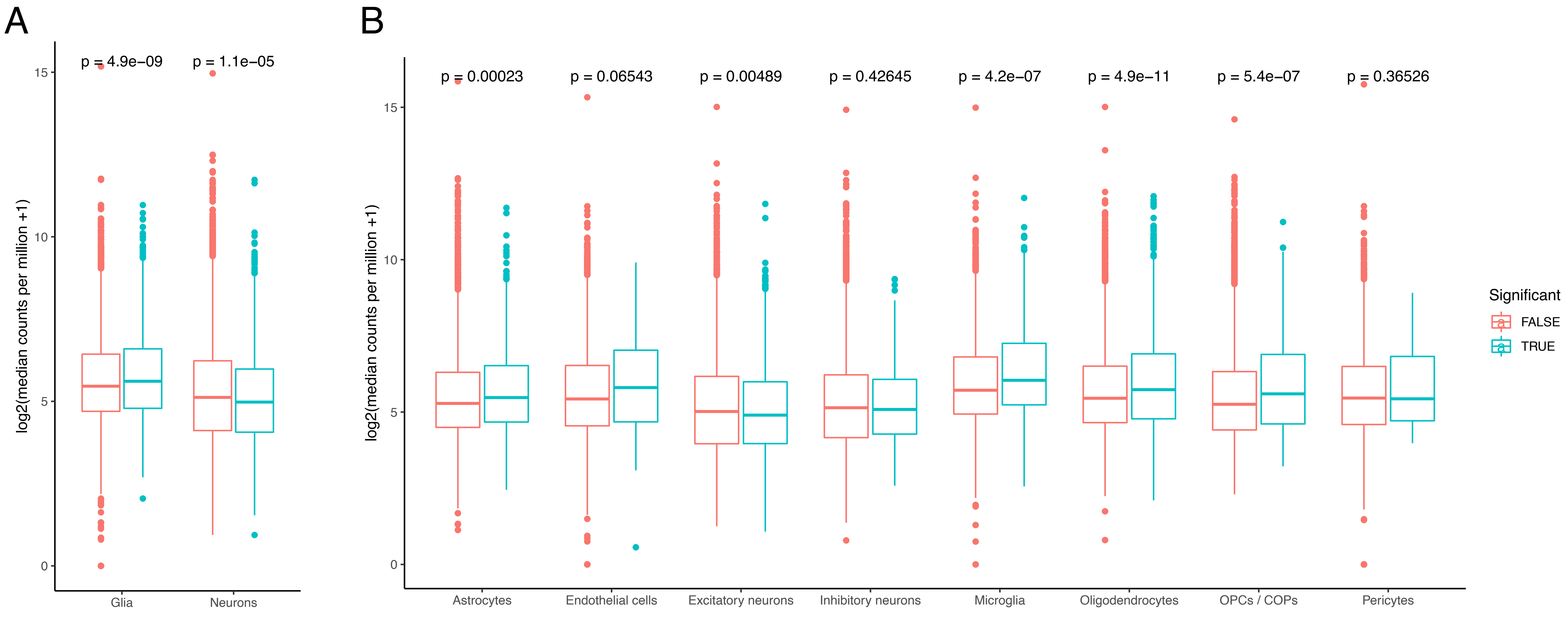
A
B
p = 4.9e−09
p = 1.1e−05
log2(median counts per million +1)
Glia
Neurons
p = 0.00023
p = 0.06543
p = 0.00489
p = 0.42645
p = 4.2e−07
p = 4.9e−11
p = 5.4e−07
p = 0.36526
Astrocytes
Endothelial cells
Excitatory neurons
Inhibitory neurons
Microglia
Oligodendrocytes
OPCs / COPs
Pericytes
Significant
FALSE
TRUE

### Figure S7

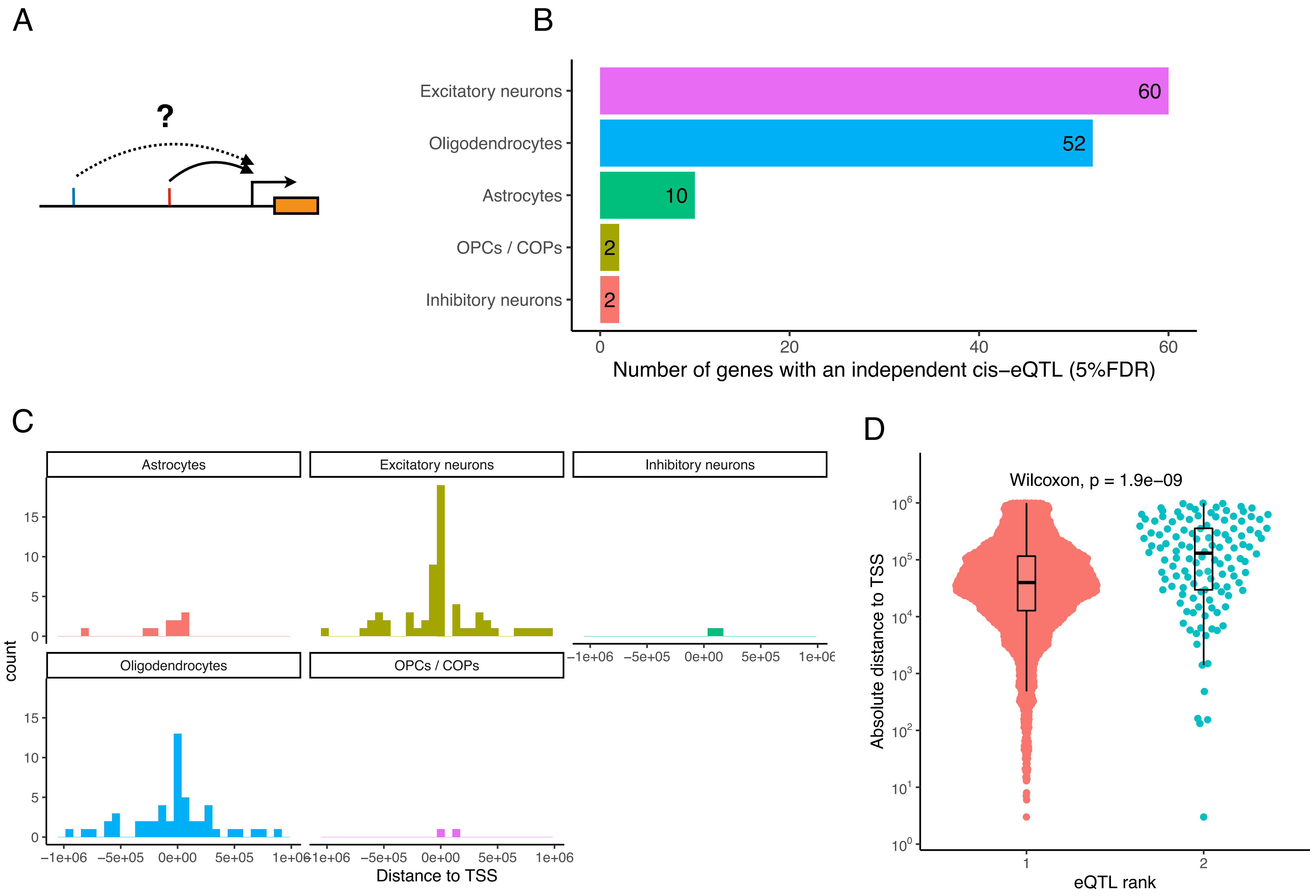

A
?
B
Excitatory neurons
Oligodendrocytes
Astrocytes
OPCs / COPs
Inhibitory neurons
60
52
10
2
2
0
20
40
60
Number of genes with an independent cis−eQTL (5%FDR)
C
Astrocytes
Excitatory neurons
Inhibitory neurons
Oligodendrocytes
OPCs / COPs
count
0
5
10
15
−1e+06
−5e+05
0e+00
5e+05
1e+06
Distance to TSS
D
Wilcoxon, p = 1.9e−09
Absolute distance to TSS
$10^0$
$10^1$
$10^2$
$10^3$
$10^4$
$10^5$
$10^6$
1
2
eQTL rank

### Figure S8

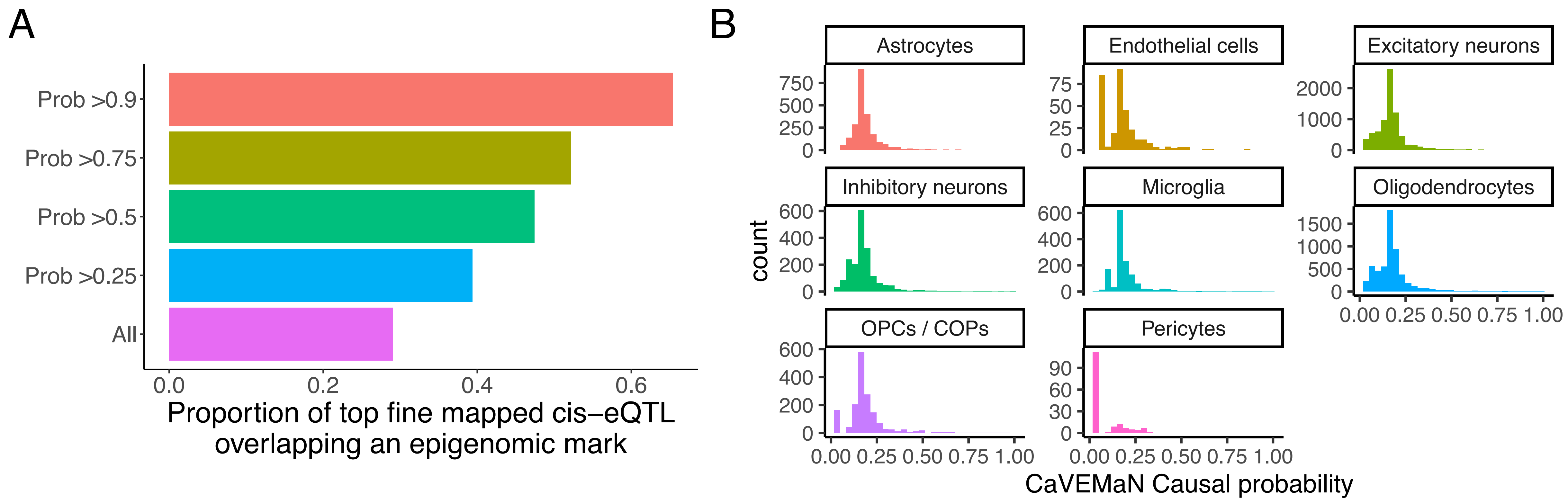

A
Prob >0.9
Prob >0.75
Prob >0.5
Prob >0.25
All
0.0
0.2
0.4
0.6
Proportion of top fine mapped cis−eQTL overlapping an epigenomic mark
B
Astrocytes
Endothelial cells
Excitatory neurons
Inhibitory neurons
Microglia
Oligodendrocytes
OPCs / COPs
Pericytes
count
0.00 0.25 0.50 0.75 1.00
CaVEMaN Causal probability

### Figure S9

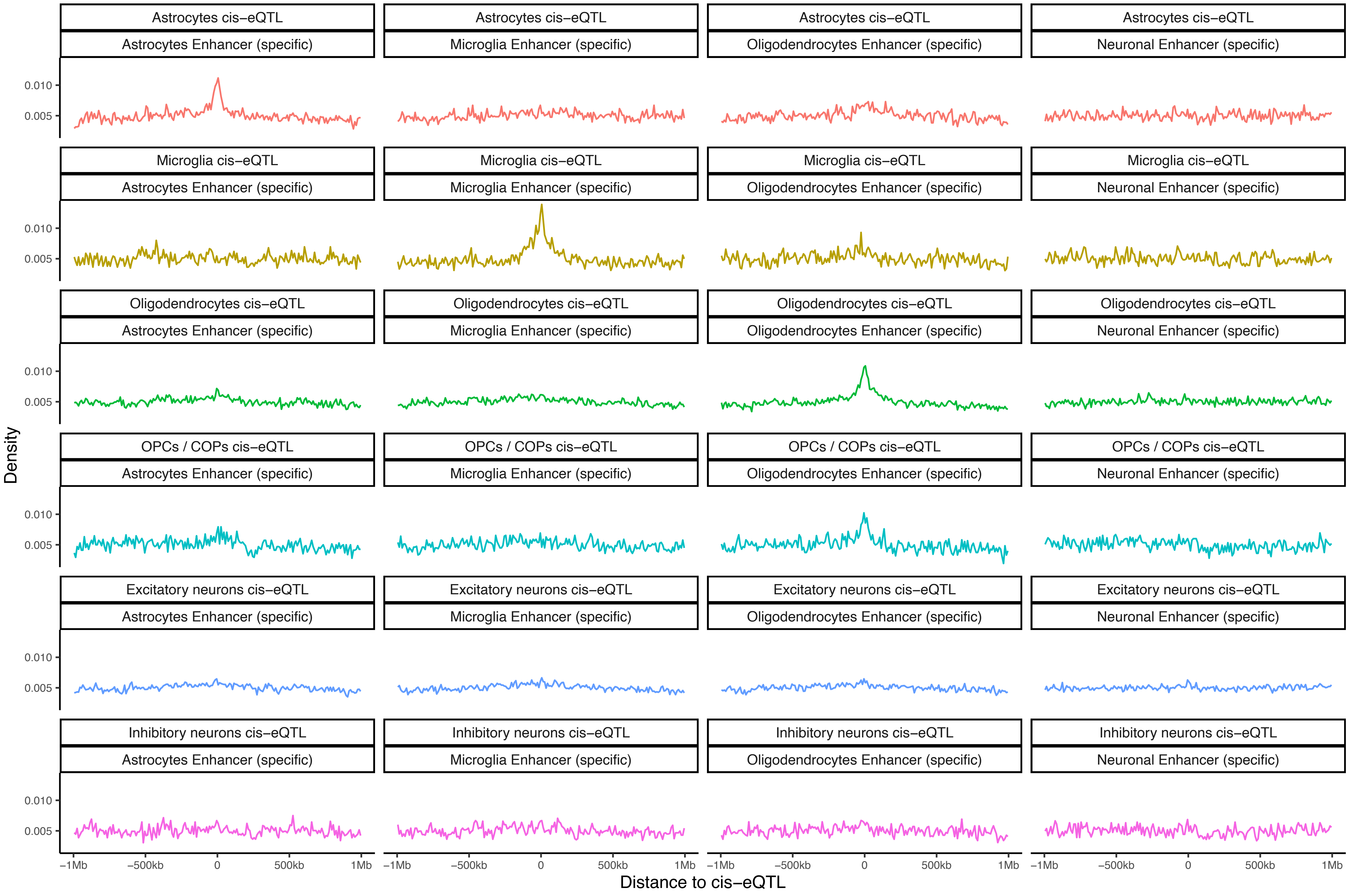
Astrocytes cis–eQTL
Microglia cis–eQTL
Oligodendrocytes cis–eQTL
OPCs / COPs cis–eQTL
Excitatory neurons cis–eQTL
Inhibitory neurons cis–eQTL
Astrocytes Enhancer (specific)
Microglia Enhancer (specific)
Oligodendrocytes Enhancer (specific)
Neuronal Enhancer (specific)
Density
0.010
0.005
–1Mb
–500kb
0
500kb
1Mb
Distance to cis–eQTL

### Figure S10

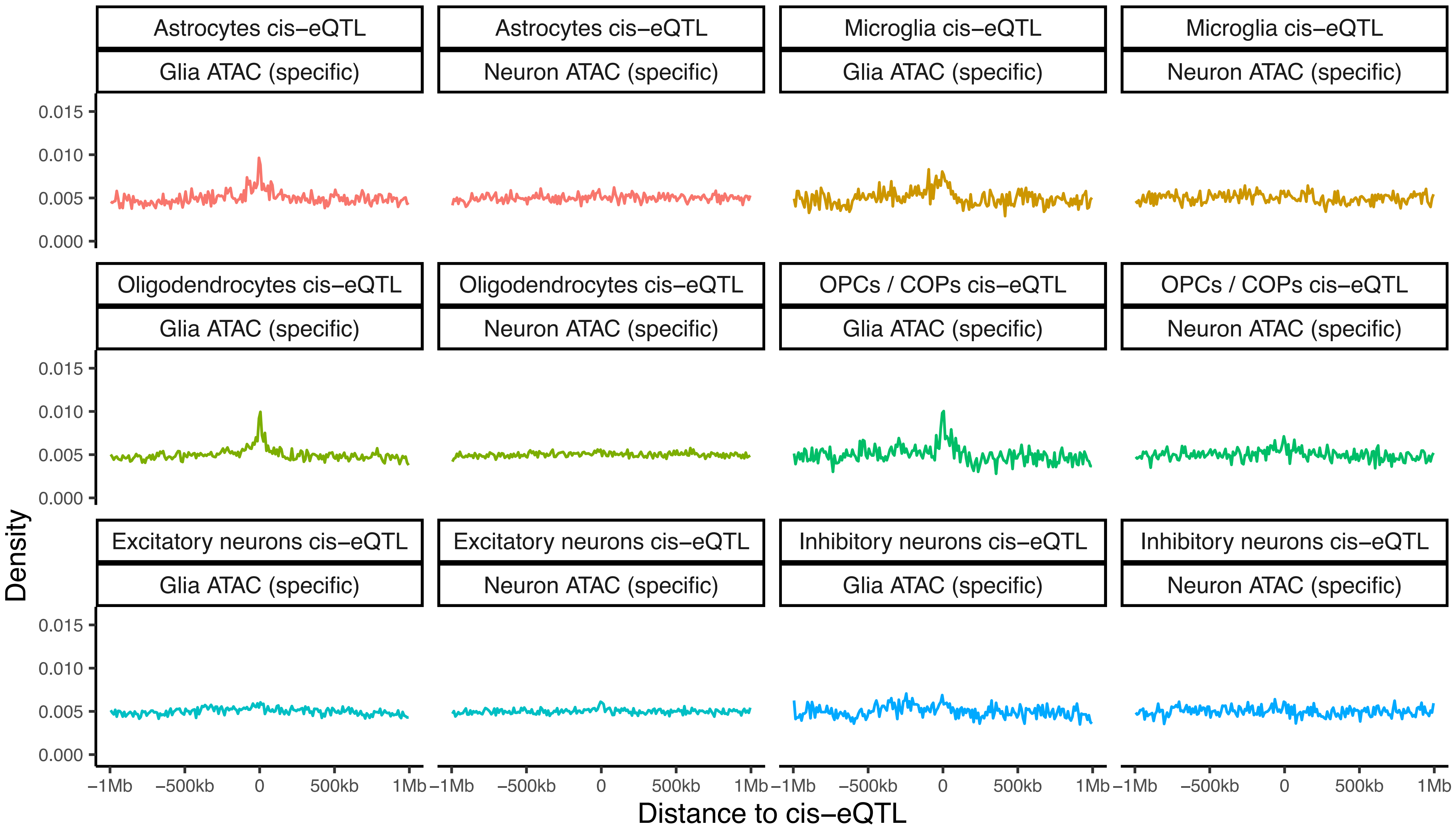
Astrocytes cis–eQTL
Glia ATAC (specific)
Astrocytes cis–eQTL
Neuron ATAC (specific)
Microglia cis–eQTL
Glia ATAC (specific)
Microglia cis–eQTL
Neuron ATAC (specific)
Oligodendrocytes cis–eQTL
Glia ATAC (specific)
Oligodendrocytes cis–eQTL
Neuron ATAC (specific)
OPCs / COPs cis–eQTL
Glia ATAC (specific)
OPCs / COPs cis–eQTL
Neuron ATAC (specific)
Excitatory neurons cis–eQTL
Glia ATAC (specific)
Excitatory neurons cis–eQTL
Neuron ATAC (specific)
Inhibitory neurons cis–eQTL
Glia ATAC (specific)
Inhibitory neurons cis–eQTL
Neuron ATAC (specific)
Density
0.015
0.010
0.005
0.000
−1Mb
−500kb
0
500kb
1Mb
Distance to cis–eQTL

### Figure S11

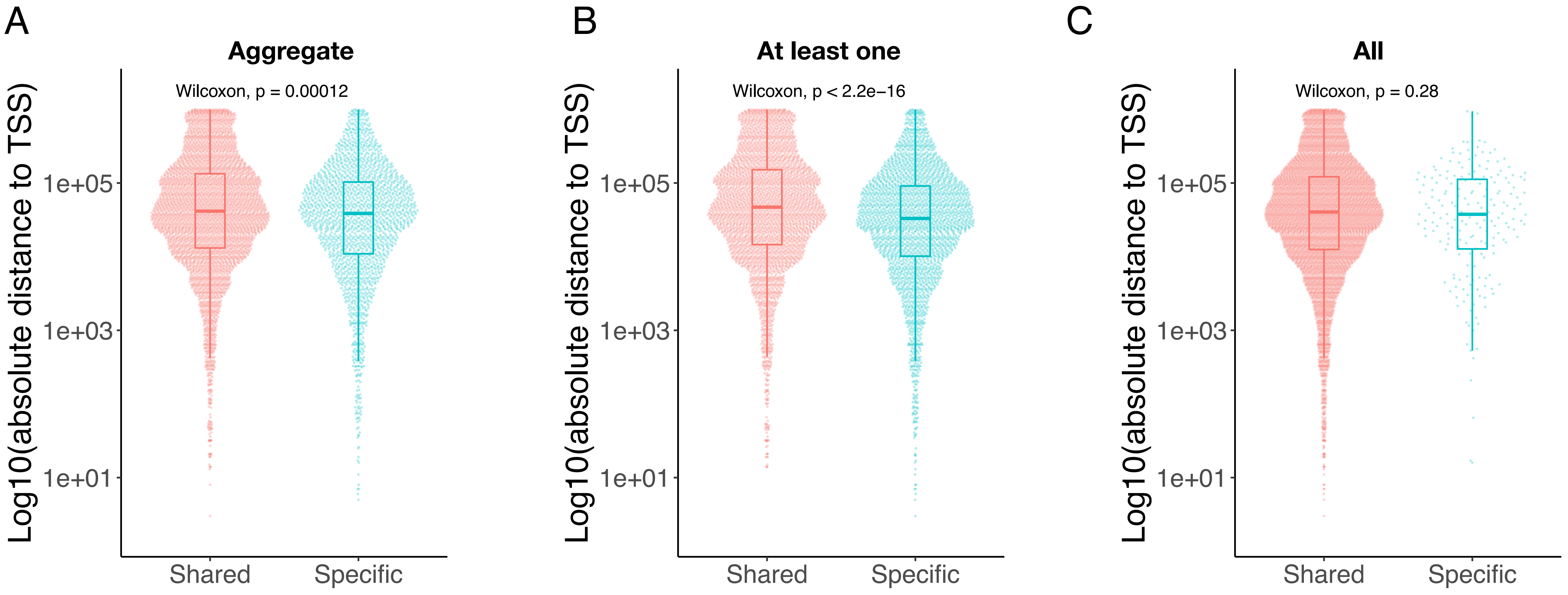
A
Aggregate
Wilcoxon, p = 0.00012
B
At least one
Wilcoxon, p < 2.2e−16
C
All
Wilcoxon, p = 0.28
Log10(absolute distance to TSS)
1e+05
1e+03
1e+01
Shared
Specific

### Figure S12

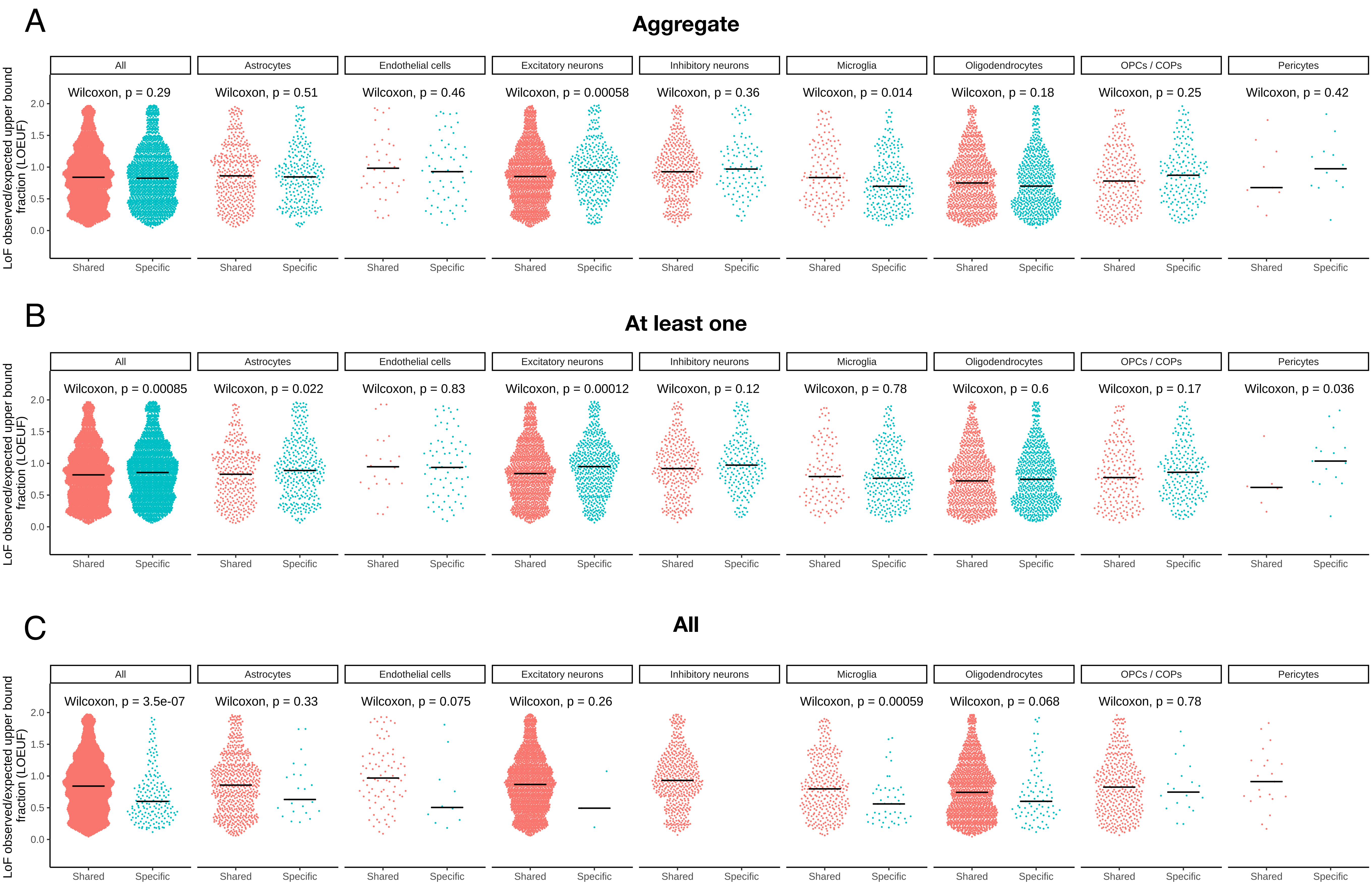

A
Aggregate
LoF observed/expected upper bound fraction (LOEUF)
2.0
1.5
1.0
0.5
0.0
All
Astrocytes
Endothelial cells
Excitatory neurons
Inhibitory neurons
Microglia
Oligodendrocytes
OPCs / COPs
Pericytes
Wilcoxon, p = 0.29
Wilcoxon, p = 0.51
Wilcoxon, p = 0.46
Wilcoxon, p = 0.00058
Wilcoxon, p = 0.36
Wilcoxon, p = 0.014
Wilcoxon, p = 0.18
Wilcoxon, p = 0.25
Wilcoxon, p = 0.42
Shared
Specific
B
At least one
LoF observed/expected upper bound fraction (LOEUF)
2.0
1.5
1.0
0.5
0.0
All
Astrocytes
Endothelial cells
Excitatory neurons
Inhibitory neurons
Microglia
Oligodendrocytes
OPCs / COPs
Pericytes
Wilcoxon, p = 0.00085
Wilcoxon, p = 0.022
Wilcoxon, p = 0.83
Wilcoxon, p = 0.00012
Wilcoxon, p = 0.12
Wilcoxon, p = 0.78
Wilcoxon, p = 0.6
Wilcoxon, p = 0.17
Wilcoxon, p = 0.036
Shared
Specific
C
All
LoF observed/expected upper bound fraction (LOEUF)
2.0
1.5
1.0
0.5
0.0
All
Astrocytes
Endothelial cells
Excitatory neurons
Inhibitory neurons
Microglia
Oligodendrocytes
OPCs / COPs
Pericytes
Wilcoxon, p = 3.5e-07
Wilcoxon, p = 0.33
Wilcoxon, p = 0.075
Wilcoxon, p = 0.26
Wilcoxon, p = 0.00059
Wilcoxon, p = 0.068
Wilcoxon, p = 0.78
Shared
Specific

### Figure S13

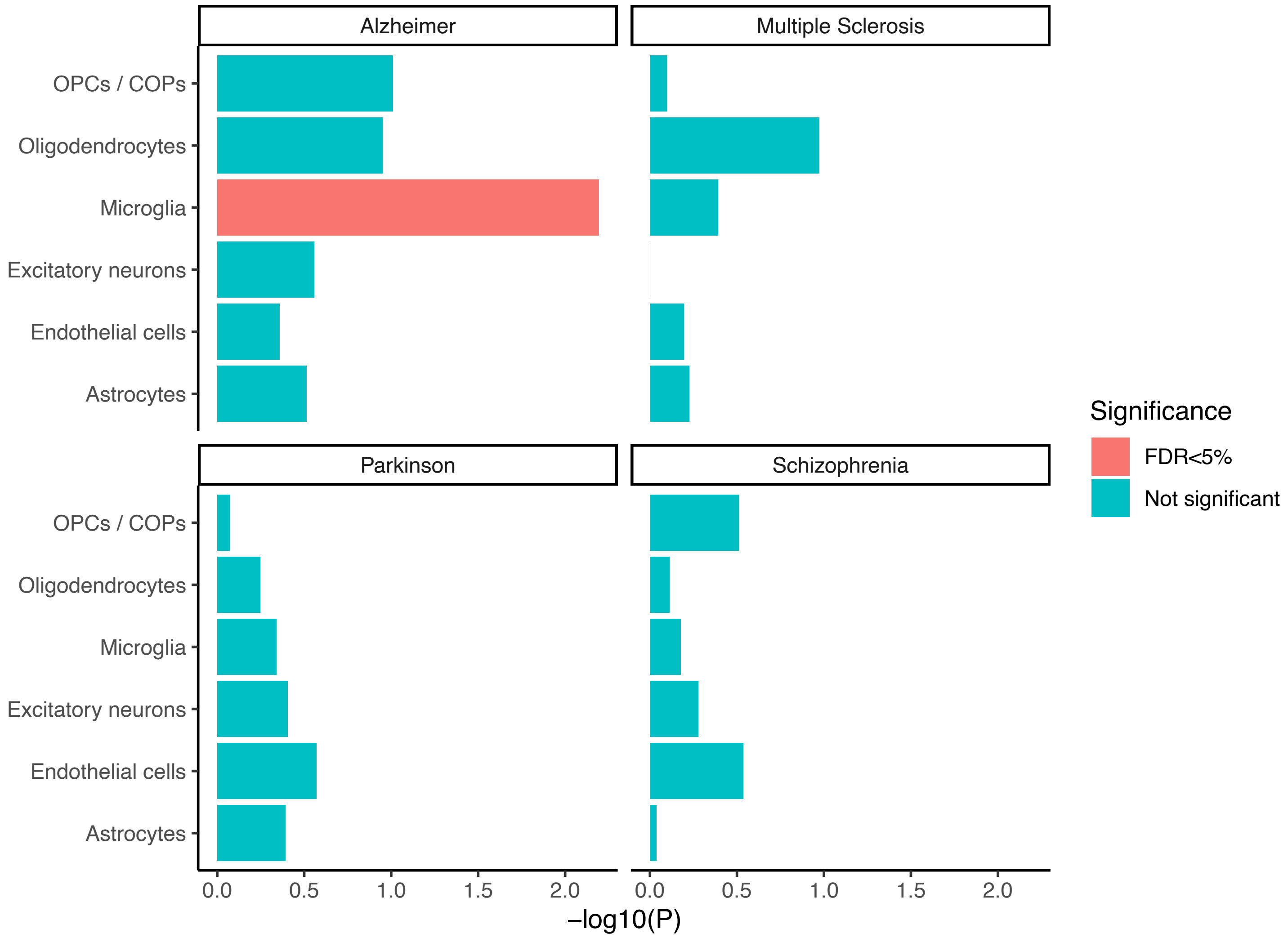

Alzheimer
Multiple Sclerosis
Parkinson
Schizophrenia
OPCs / COPs
Oligodendrocytes
Microglia
Excitatory neurons
Endothelial cells
Astrocytes
0.0
0.5
1.0
1.5
2.0
–log10(P)
Significance
FDR<5%
Not significant

### Figure S14

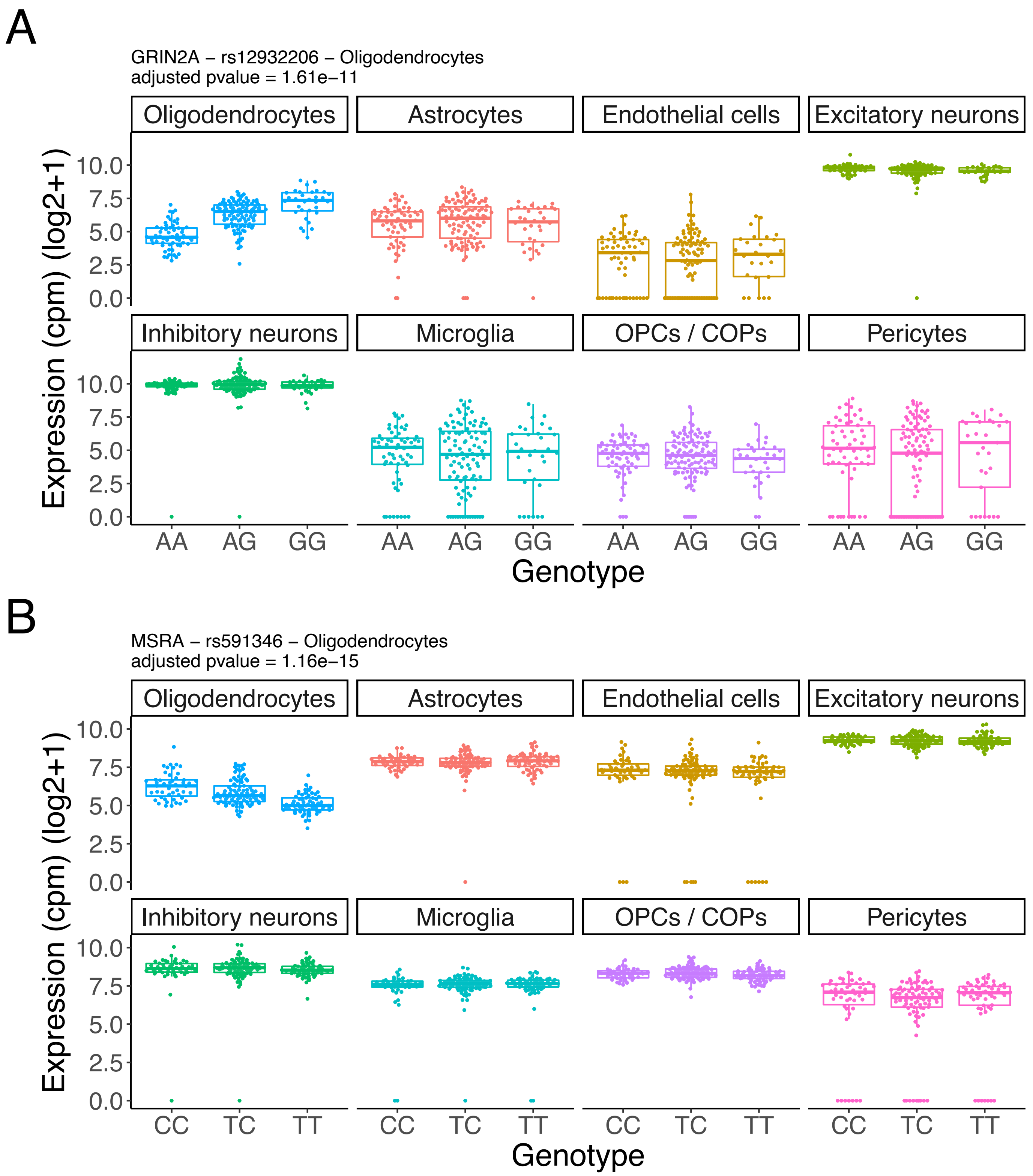
A
GRIN2A – rs12932206 – Oligodendrocytes
adjusted pvalue = 1.61e−11
Oligodendrocytes
Astrocytes
Endothelial cells
Excitatory neurons
Inhibitory neurons
Microglia
OPCs / COPs
Pericytes
Expression (cpm) (log2+1)
10.0
7.5
5.0
2.5
0.0
AA AG GG
Genotype
B
MSRA – rs591346 – Oligodendrocytes
adjusted pvalue = 1.16e−15
Oligodendrocytes
Astrocytes
Endothelial cells
Excitatory neurons
Inhibitory neurons
Microglia
OPCs / COPs
Pericytes
Expression (cpm) (log2+1)
10.0
7.5
5.0
2.5
0.0
CC TC TT
Genotype

### Figure S15

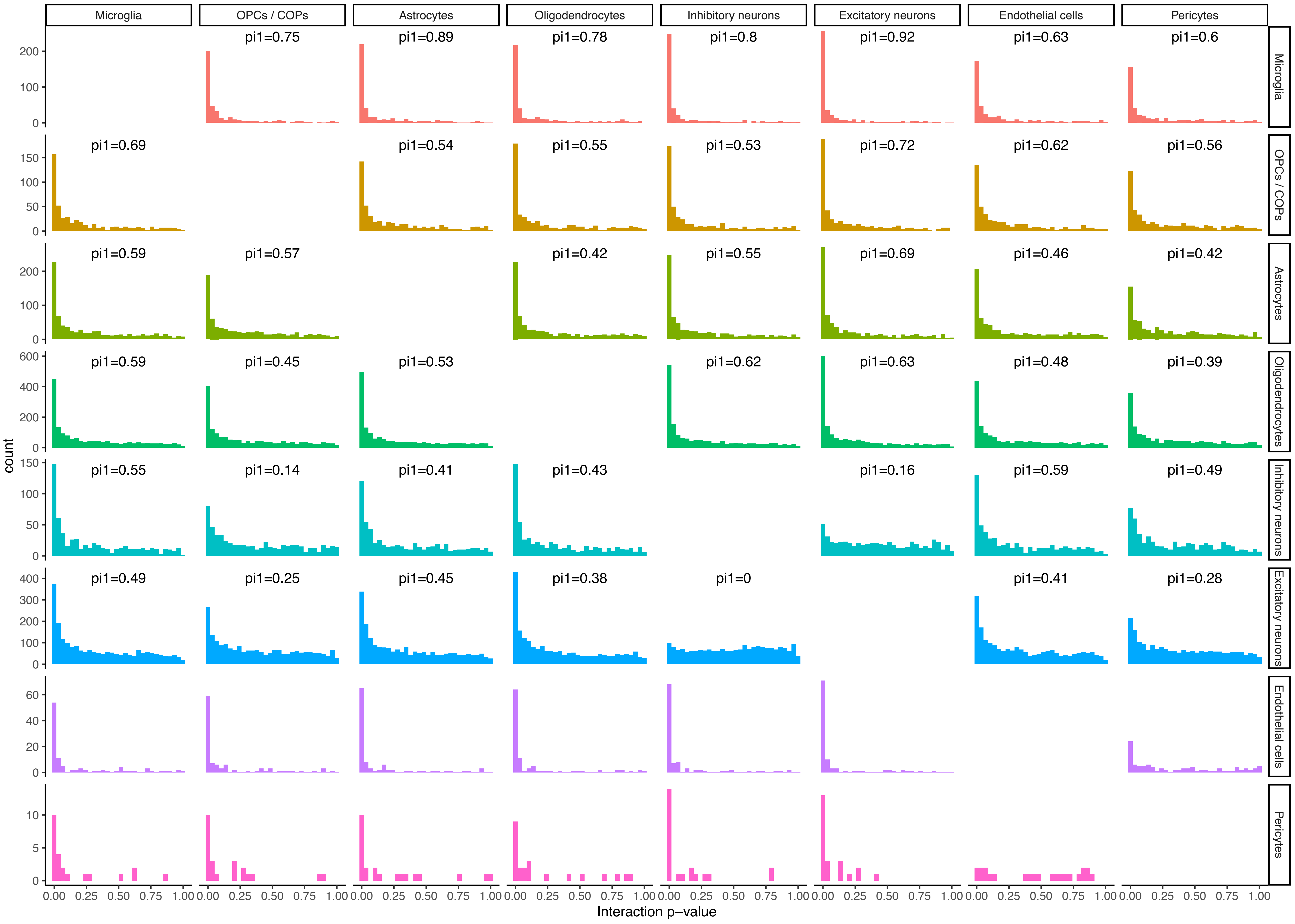

Microglia
OPCs / COPs
Astrocytes
Oligodendrocytes
Inhibitory neurons
Excitatory neurons
Endothelial cells
Pericytes
pi1=0.75
pi1=0.89
pi1=0.78
pi1=0.8
pi1=0.92
pi1=0.63
pi1=0.6
pi1=0.69
pi1=0.54
pi1=0.55
pi1=0.53
pi1=0.72
pi1=0.62
pi1=0.56
pi1=0.59
pi1=0.57
pi1=0.42
pi1=0.55
pi1=0.69
pi1=0.46
pi1=0.42
pi1=0.59
pi1=0.45
pi1=0.53
pi1=0.62
pi1=0.63
pi1=0.48
pi1=0.39
pi1=0.55
pi1=0.14
pi1=0.41
pi1=0.43
pi1=0.16
pi1=0.59
pi1=0.49
pi1=0.49
pi1=0.25
pi1=0.45
pi1=0.38
pi1=0
pi1=0.41
pi1=0.28
count
Interaction p−value
0.00 0.25 0.50 0.75 1.00

### Figure S16

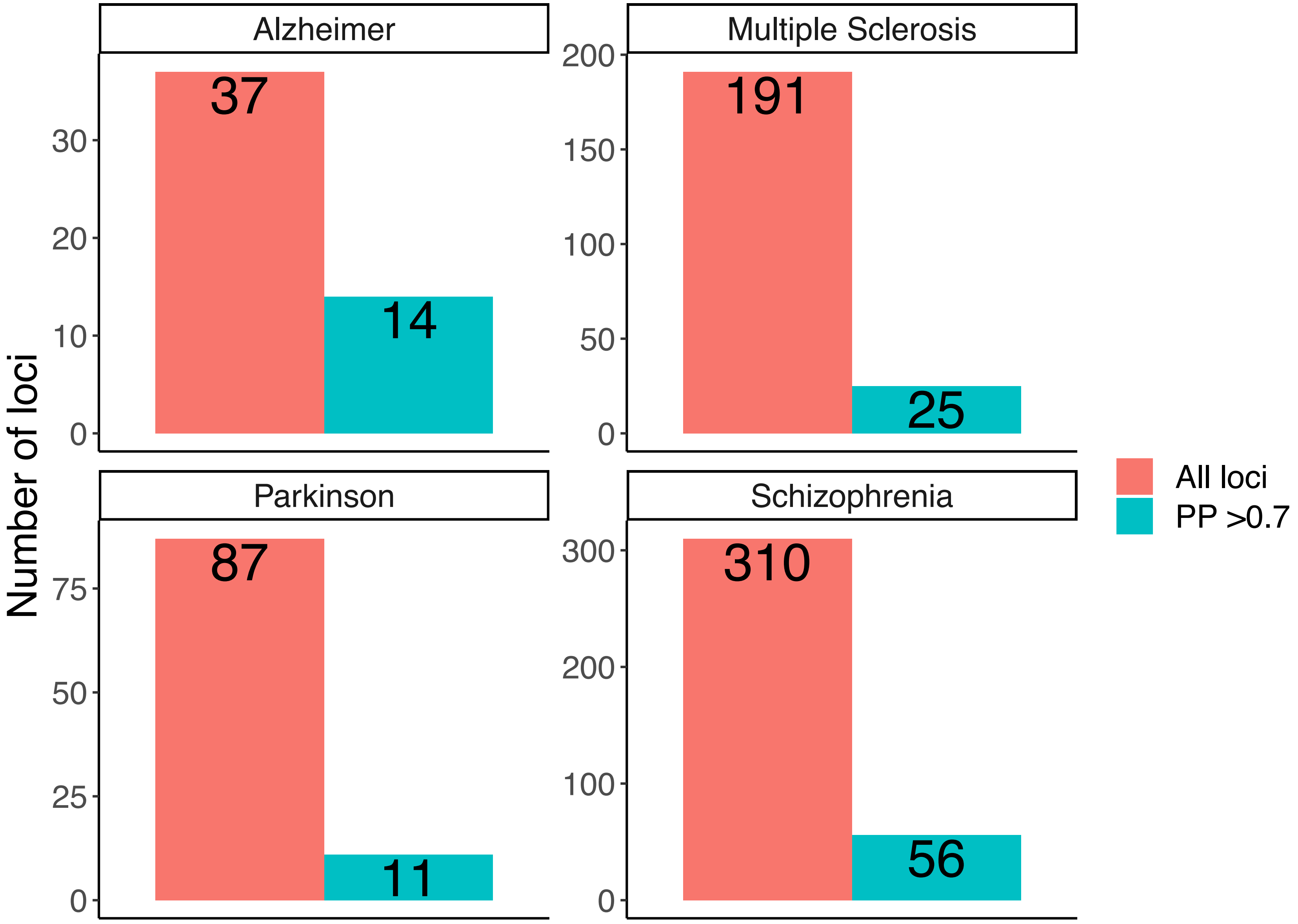
Alzheimer
Multiple Sclerosis
Parkinson
Schizophrenia
Number of loci
37
14
191
25
87
11
310
56
30
20
10
0
200
150
100
50
0
75
50
25
0
300
200
100
0
All loci
PP >0.7

### Figure S17

| | Microglia | Oligodendrocytes | Endothelial cells | Astrocytes | Excitatory neurons | Inhibitory neurons | Pericytes | OPCs / COPs |
|---|---|---|---|---|---|---|---|---|
| APH1B | 5.48 | 5.19 | 4.33 | 5.32 | 4.51 | 4.97 | 5.10 | 5.38 |
| BIN1 | 8.56 | 8.91 | 5.22 | 5.11 | 6.51 | 6.51 | 5.48 | 5.41 |
| CASS4 | 5.78 | 1.78 | 6.02 | 2.29 | 1.72 | 1.64 | 3.05 | 1.29 |
| CD2AP | 7.13 | 4.83 | 7.45 | 5.50 | 5.34 | 5.49 | 6.32 | 4.87 |
| SIGLEC9 | 5.60 | 0.13 | 0.00 | 0.22 | 0.21 | 0.00 | 0.00 | 0.00 |
| CLU | 6.47 | 7.22 | 8.38 | 10.96 | 8.63 | 7.90 | 8.26 | 7.69 |
| CR1 | 2.69 | 4.24 | 0.00 | 0.84 | 0.63 | 0.41 | 0.00 | 1.04 |
| PFKFB2 | 5.03 | 3.76 | 2.57 | 6.16 | 4.20 | 3.77 | 2.91 | 2.94 |
| USP6NL | 8.20 | 3.96 | 8.18 | 5.65 | 3.95 | 4.72 | 6.58 | 5.96 |
| ZYX | 4.71 | 2.79 | 4.97 | 3.65 | 5.08 | 4.43 | 5.05 | 3.56 |
| INPP5D | 9.82 | 1.47 | 7.98 | 3.58 | 2.41 | 1.04 | 5.25 | 1.16 |
| PICALM | 10.22 | 9.54 | 9.57 | 7.78 | 7.35 | 7.30 | 8.73 | 7.94 |
| RABEP1 | 7.77 | 7.33 | 7.00 | 6.86 | 7.36 | 7.24 | 7.11 | 6.79 |
| RIN3 | 8.83 | 1.43 | 4.90 | 4.56 | 3.28 | 2.91 | 3.80 | 5.04 |
| TREM2 | 6.33 | 0.22 | 0.00 | 0.06 | 0.04 | 0.00 | 0.00 | 0.00 |

### Figure S18

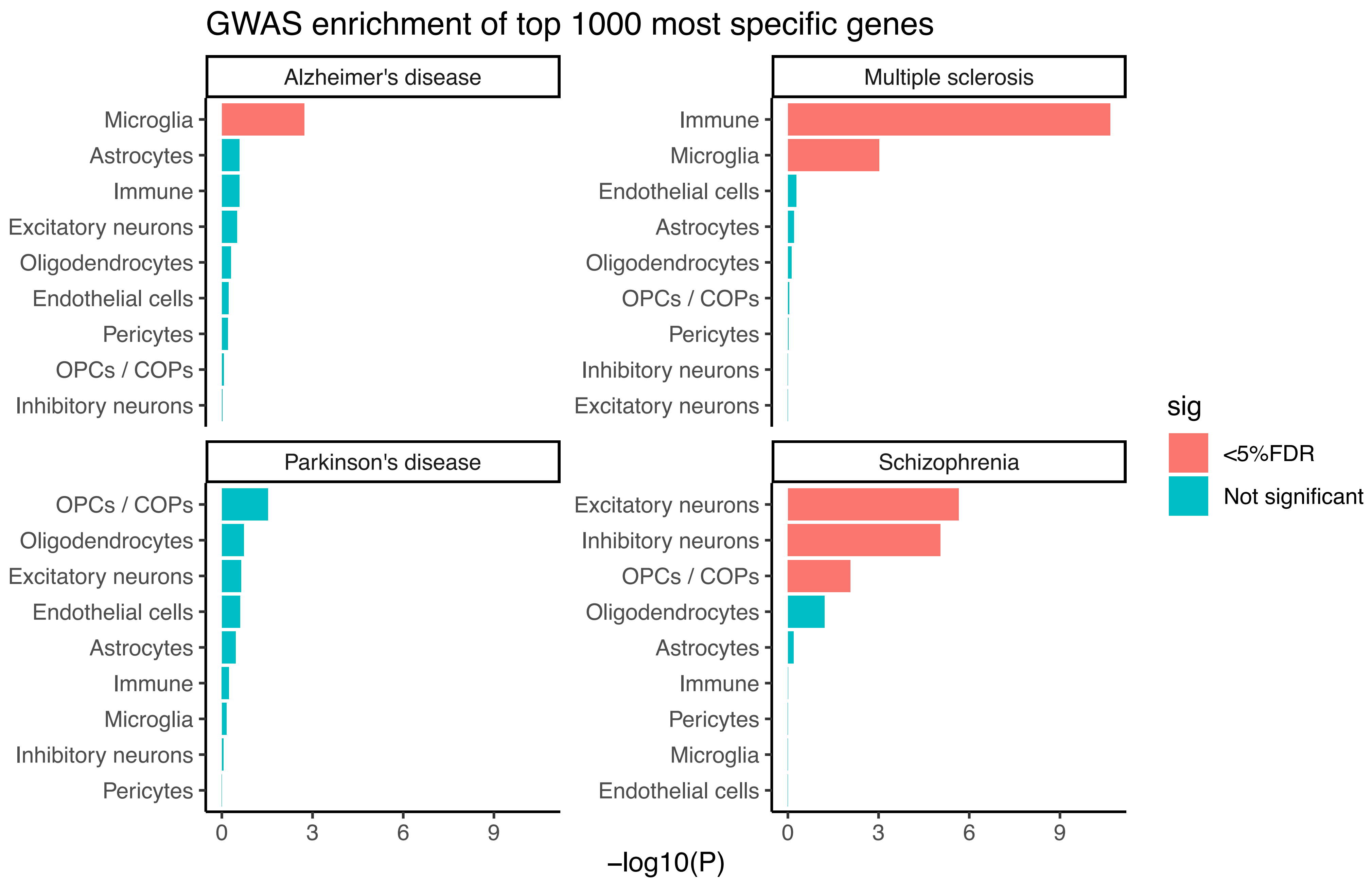

GWAS enrichment of top 1000 most specific genes
Alzheimer's disease
Microglia
Astrocytes
Immune
Excitatory neurons
Oligodendrocytes
Endothelial cells
Pericytes
OPCs / COPs
Inhibitory neurons
Multiple sclerosis
Immune
Microglia
Endothelial cells
Astrocytes
Oligodendrocytes
OPCs / COPs
Pericytes
Inhibitory neurons
Excitatory neurons
Parkinson's disease
OPCs / COPs
Oligodendrocytes
Excitatory neurons
Endothelial cells
Astrocytes
Immune
Microglia
Inhibitory neurons
Pericytes
Schizophrenia
Excitatory neurons
Inhibitory neurons
OPCs / COPs
Oligodendrocytes
Astrocytes
Immune
Pericytes
Microglia
Endothelial cells
0
3
6
9
0
3
6
9
−log10(P)
sig
<5%FDR
Not significant

### Figure S19

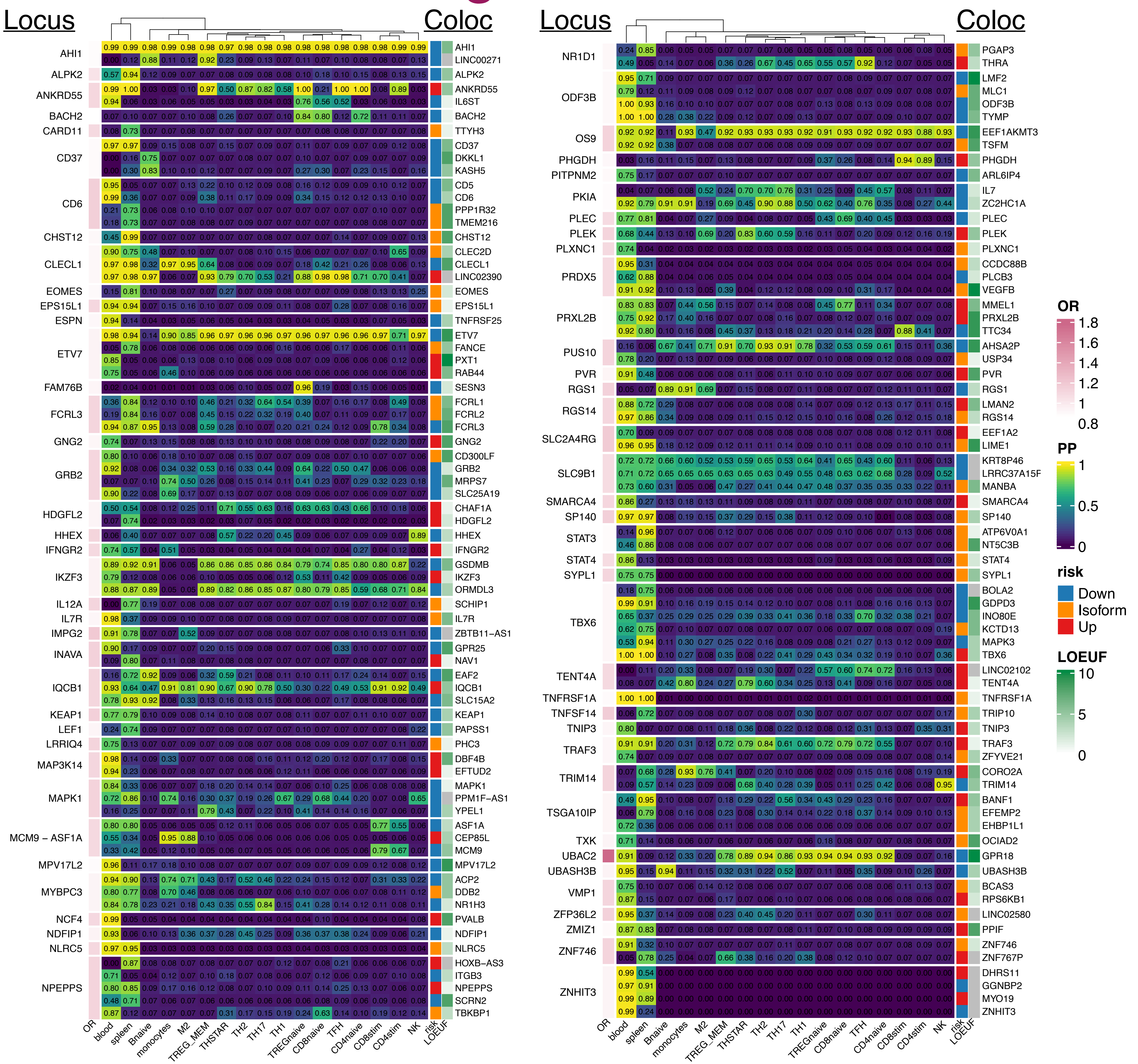
Locus
Coloc
AHI1
ALPK2
ANKRD55
BACH2
CARD11
CD37
CD6
CHST12
CLECL1
EOMES
EPS15L1
ESPN
ETV7
FAM76B
FCRL3
GNG2
GRB2
HDGFL2
HHEX
IFNGR2
IKZF3
IL12A
IL7R
IMPG2
INAVA
IQCB1
KEAP1
LEF1
LRRIQ4
MAP3K14
MAPK1
MCM9 – ASF1A
MPV17L2
MYBPC3
NCF4
NDFIP1
NLRC5
NPEPPS
AHI1
LINC00271
ALPK2
ANKRD55
IL6ST
BACH2
TTYH3
CD37
DKKL1
KASH5
CD5
CD6
PPP1R32
TMEM216
CHST12
CLEC2D
CLECL1
LINC02390
EOMES
EPS15L1
TNFRSF25
ETV7
FANCE
PXT1
RAB44
SESN3
FCRL1
FCRL2
FCRL3
GNG2
CD300LF
GRB2
MRPS7
SLC25A19
CHAF1A
HDGFL2
HHEX
IFNGR2
GSDMB
IKZF3
ORMDL3
SCHIP1
IL7R
ZBTB11-AS1
GPR25
NAV1
EAF2
IQCB1
SLC15A2
KEAP1
PAPSS1
PHC3
DBF4B
EFTUD2
MAPK1
PPM1F-AS1
YPEL1
ASF1A
CEP85L
MCM9
MPV17L2
ACP2
DDB2
NR1H3
PVALB
NDFIP1
NLRC5
HOXB-AS3
ITGB3
NPEPPS
SCRN2
TBKBP1
NR1D1
ODF3B
OS9
PHGDH
PITPNM2
PKIA
PLEC
PLEK
PLXNC1
PRDX5
PRXL2B
PUS10
PVR
RGS1
RGS14
SLC2A4RG
SLC9B1
SMARCA4
SP140
STAT3
STAT4
SYPL1
TBX6
TENT4A
TNFRSF1A
TNFSF14
TNIP3
TRAF3
TRIM14
TSGA10IP
TXK
UBAC2
UBASH3B
VMP1
ZFP36L2
ZMIZ1
ZNF746
ZNHIT3
PGAP3
THRA
LMF2
MLC1
ODF3B
TYMP
EEF1AKMT3
TSFM
PHGDH
ARL6IP4
IL7
ZC2HC1A
PLEC
PLEK
PLXNC1
CCDC88B
PLCB3
VEGFB
MMEL1
PRXL2B
TTC34
AHSA2P
USP34
PVR
RGS1
LMAN2
RGS14
EEF1A2
LIME1
KRT8P46
LRRC37A15F
MANBA
SMARCA4
SP140
ATP6V0A1
NT5C3B
STAT4
SYPL1
BOLA2
GDPD3
INO80E
KCTD13
MAPK3
TBX6
LINC02102
TENT4A
TNFRSF1A
TRIP10
TNIP3
TRAF3
ZFYVE21
CORO2A
TRIM14
BANF1
EFEMP2
EHBP1L1
OCIAD2
GPR18
UBASH3B
BCAS3
RPS6KB1
LINC02580
PPIF
ZNF746
ZNF767P
DHRS11
GGNBP2
MYO19
ZNHIT3
OR
blood
spleen
Bnaive
monocytes
M2
TREG_MEM
THSTAR
TH2
TH17
TH1
TREGnaive
CD8naive
TFH
CD4naive
CD8stim
CD4stim
NK
risk
LOEUF
OR
1.8
1.6
1.4
1.2
1
0.8
PP
1
0.5
0
risk
Down
Isoform
Up
LOEUF
10
5
0

### Figure S20

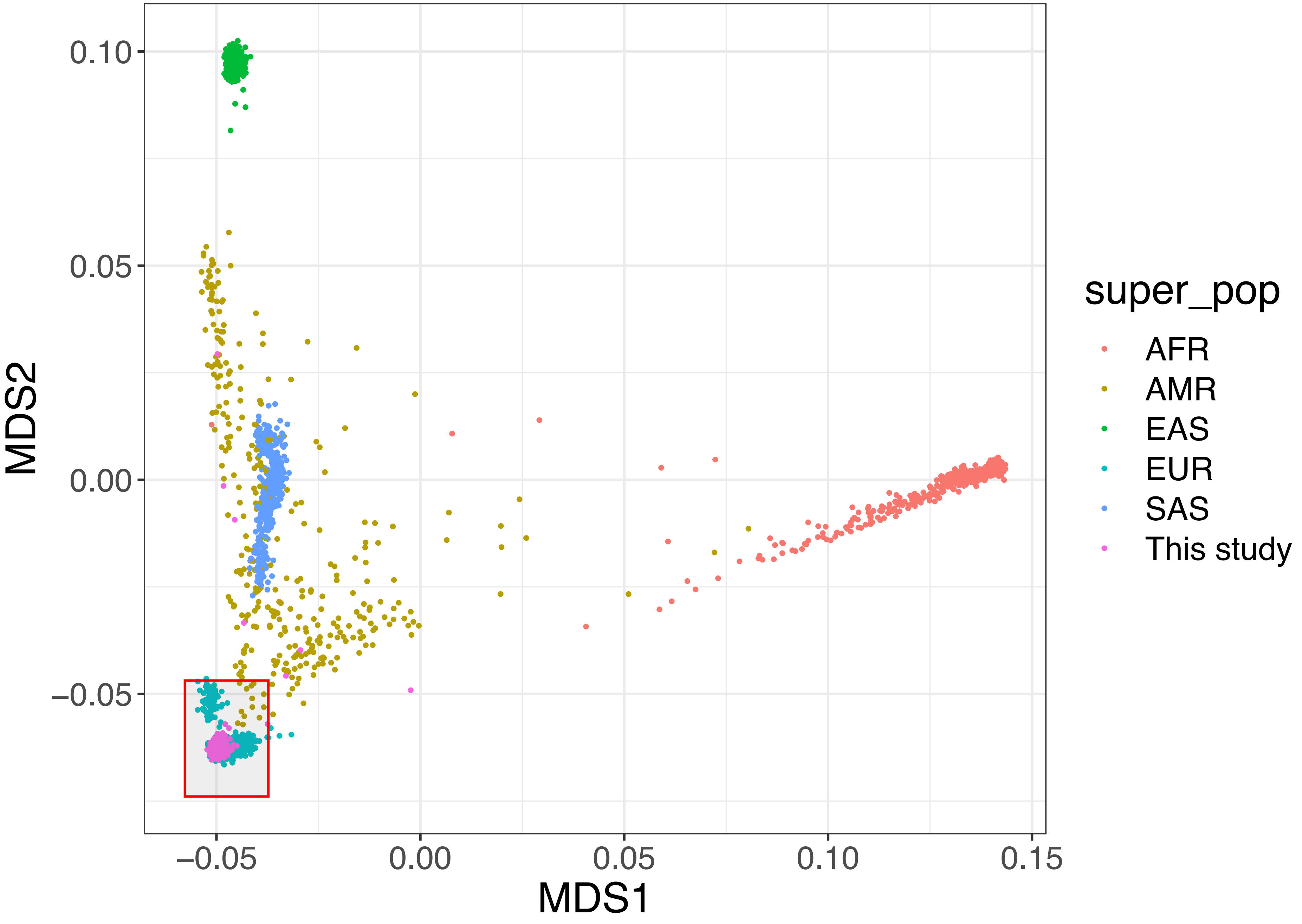

0.10
0.05
0.00
−0.05
MDS2
−0.05
0.00
0.05
0.10
0.15
MDS1
super_pop
AFR
AMR
EAS
EUR
SAS
This study

### Figure S21

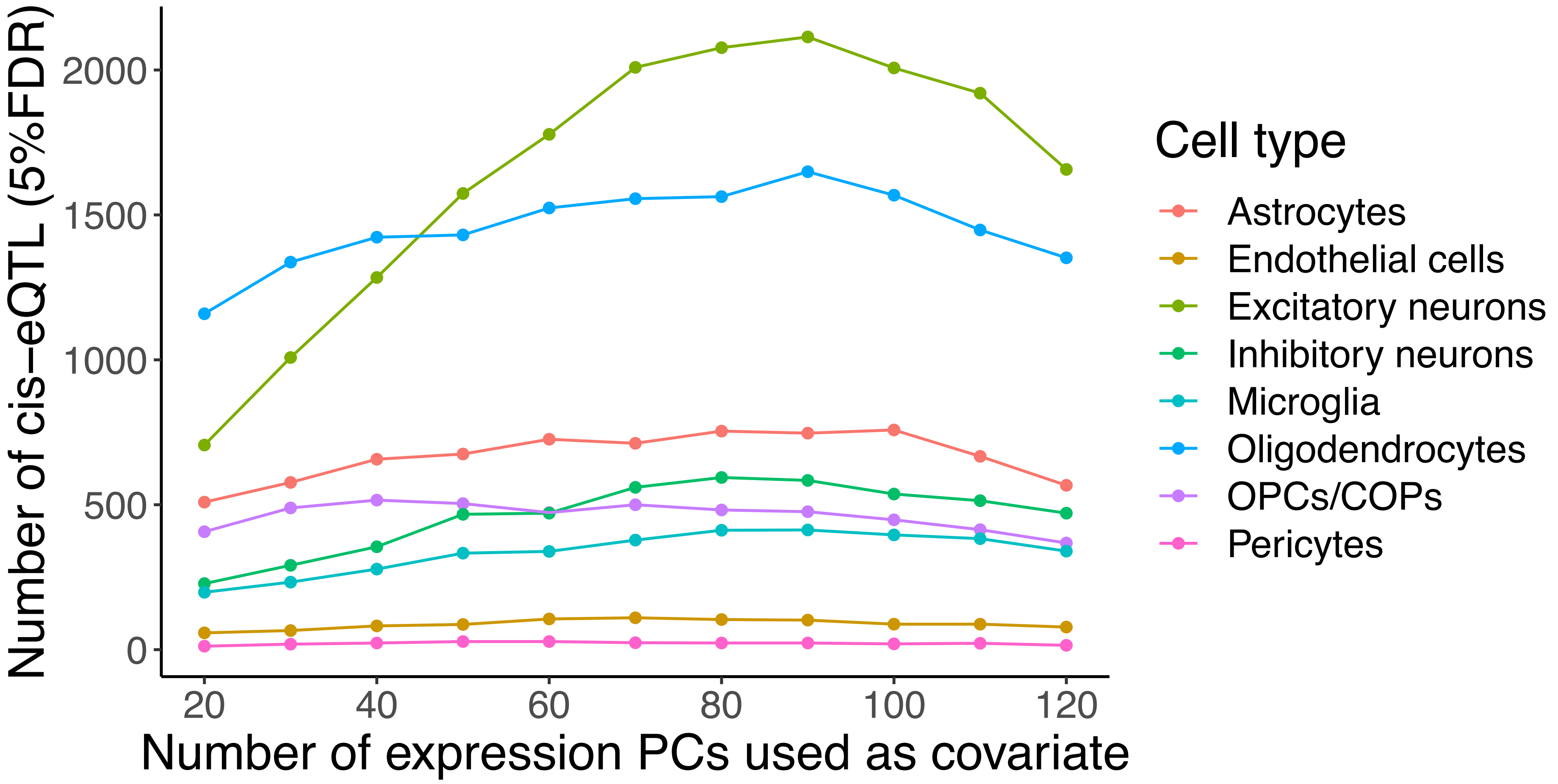

Number of cis−eQTL (5%FDR)
2000
1500
1000
500
0
20
40
60
80
100
120
Number of expression PCs used as covariate
Cell type
Astrocytes
Endothelial cells
Excitatory neurons
Inhibitory neurons
Microglia
Oligodendrocytes
OPCs/COPs
Pericytes
